# SAFETY, PHARMACOKINETICS, AND PHARMACODYNAMICS OF A CLC-1 INHIBITOR - A FIRST-IN-CLASS COMPOUND THAT ENHANCES MUSCLE EXCITABILITY: A PHASE I, SINGLE- AND MULTIPLE-ASCENDING DOSE STUDY

**DOI:** 10.1101/2024.02.15.24302858

**Authors:** Titia Q. Ruijs, Catherine M.K.E. de Cuba, Jules A.A.C. Heuberger, John Hutchison, Jane Bold, Thomas S. Grønnebæk, Klaus G Jensen, Eva Chin, Jorge A. Quiroz, Thomas K. Petersen, Peter Flagstad, Marieke L. de Kam, Michiel J. van Esdonk, Erica Klaassen, Robert J. Doll, Ingrid W. Koopmans, Annika A. de Goede, Thomas H. Pedersen, Geert Jan Groeneveld

## Abstract

NMD670 is a first-in-class inhibitor of skeletal muscle-specific chloride channel ClC-1, developed to improve muscle weakness and fatigue in neuromuscular diseases. Preclinical studies show that ClC-1 inhibition enhances muscle excitability, improving muscle contractility and strength. We describe the first-in-human administration of ClC-1 inhibitor NMD670. In this randomized, double-blind, placebo-controlled study we evaluated safety, pharmacokinetics, and pharmacodynamics of single and multiple doses of NMD670 in healthy male and female subjects. Single-ascending doses were administered in a (partial) cross-over design; multiple-ascending doses were administered in a parallel design. Differences in pharmacokinetics between males/females and fed/fasted state were evaluated. Pharmacodynamic effects were evaluated using muscle velocity recovery cycles (MVRC), and analyzed using mixed effects modelling, with baseline as covariate. NMD670 was considered safe and well-tolerated. Symptoms of myotonia were observed at the highest dose levels. Moreover, NMD670 significantly increased the following MVRC parameters after a single dose of 1200mg compared to placebo: early supernormality (estimated difference (ED) 2.04; 95% confidence interval (CI) (0.379, 3.70); p=0.0242); early supernormality after 5 conditioning stimuli (ED 2.51; 95%CI (0.599, 4.41); p=0.0177; supernormality at 20 ms (ED 2.78; 95%CI (1.377, 4.181); p=0.0021. Importantly, the results of this study indicate pharmacological target engagement of NMD670 as ClC-1 inhibitor, at dose levels that were considered safe in healthy subjects. Firstly, because myotonia was an expected exaggerated on-target pharmacological effect. Secondly, because the effects on MVRC indicate increased muscle cell excitability. This study in healthy subjects indicates proof-of-mechanism and provides a solid base for translation to patients with neuromuscular diseases.

## Introduction

Skeletal muscle-specific chloride channel ClC-1 plays a major role in the regulation of muscle cell excitability. In the resting muscle, ClC-1 provides an inhibitory current responsible for 80% of the resting muscle membrane conductance ^1^. Upon muscle activation, the extracellular potassium concentration increases, and the number of active sodium channels decreases, compromising muscle activation. To maintain proper muscle function, the inhibitory ClC-1 current is decreased by cellular signals that arise during muscle activity including activation of Protein Kinase C. This endogenous ClC-1 inhibition during muscle activity enhances neuromuscular transmission strength. With prolonged, intense muscle activity, however, there is reversed regulation: the inhibitory ClC-1 current increases largely, possibly to protect muscles with decreased cellular energy levels ^2^.

Neuromuscular diseases (NMDs) are a variable group of disorders with different pathologies, but most NMDs lead to muscle weakness and major disability. In a selection of NMDs, such as myasthenia gravis (MG) and Lambert-Eaton myasthenic syndrome (LEMS), muscle weakness is caused by a failure of muscle activation due to compromised neuromuscular transmission ^3^. In preclinical models of MG, inhibition of ClC-1 has been shown to improve neuromuscular function ^4^. Taken together, the improved muscle excitability caused by endogenous ClC-1 inhibition during intense muscle activity, and the recovery of muscle function in pre-clinical MG models with ClC-1 inhibition, suggest that inhibition of ClC-1 could be an interesting novel target for certain NMDs, such as MG.

In this study we evaluated the effects of NMD670 – a first-in-class compound that selectively inhibits ClC-1 – when administered to humans for the first time. The aim of this study was to evaluate the safety, pharmacokinetics (PK) and pharmacodynamics (PD) of single-ascending doses (SAD) and multiple-ascending doses (MAD) of NMD670 in healthy male and female subjects. After this work, we have demonstrated effects of NMD670 in patients with MG (unpublished data).

## Methods

The study was approved by Ethics Committee Stichting “Beoordeling Ethiek Biomedisch Onderzoek”, The Netherlands, and registered in NTR: NL8692. The study was conducted in accordance with the Declaration of Helsinki and International Conference on Harmonization Good Clinical Practice.

### Subjects

Healthy male (18-45 years) and female subjects of non-childbearing potential (18-65 years) were included in the SAD study. The MAD study included healthy male subjects (18-65 years). Health status was confirmed during a medical screening, which involved evaluation of medical history, physical examination, electrocardiogram (ECG), vital signs, and laboratory tests. Body mass index (BMI) was 18-30 kg/m^2^, with minimum weight 50 kg. Subjects who smoked >10 cigarettes per day were excluded; use of nicotine was prohibited during study visits. Subjects with history of illicit drug- or alcohol abuse, or a positive drug/alcohol test, were excluded. Subject were asked to adhere to certain lifestyle restrictions. Use of medication, dietary supplements, CYP450 isoenzyme modulating products, alcohol, illicit drugs, caffeine, and nicotine was not allowed. Excessive exercise was prohibited for seven days pre-dose. The nutritional composition of meals and dining times were standardized during inpatient periods. In the SAD study, subjects were asked to fast from the night before dosing until three hours post-dose, with exception of the investigation of food effect. In the MAD study, subjects were not fasted, because no food effect was found in the SAD study. Strong sun exposure was to be avoided and subjects were asked to use contraception.

### Study design

The study was performed between July 2020 and August 2021 at the Centre for Human Drug Research, Leiden, NL. The study design is summarized in Figure S1. The SAD study was a randomized, double-blind, placebo-controlled partial cross-over study. A partial cross-over design was chosen to allow for intraindividual comparison of PD endpoints. Subjects were divided in three cohorts: each cohort consisted of nine subjects who each had three study sessions. Subjects within cohorts were randomized 6:3 (NMD670:placebo). Each subject received NMD670 on two visits and placebo on one visit, the order of the treatments placebo and rising doses of NMD670 was randomized. For dose escalation between cohorts, blinded interim data was reviewed for PK, safety and PD. Cohort 2 returned for an additional fourth visit to evaluate food effect, during which subjects received 800 mg NMD670 after a high-fat, high calorie breakfast, in the same randomization as the fasted condition at the same dose level.

There was a wash-out of at least 10 days between treatments. The SAD study included a female cohort to evaluate gender effects in which subjects were randomized to receive a single dose of 800 mg NMD670 or placebo (6:2). The MAD study had a randomized, double-blind, placebo-controlled, parallel study design. Subjects were randomized to receive either NMD670 or placebo (6:2) for a duration of ten consecutive days. Both the SAD and MAD study employed sentinel dosing for the first dose of the first cohort.

There were two important changes in study conduct during study execution. The study was put on temporary halt because of the occurrence of moderately severe myotonia after administration of NMD670 1600 mg in one subject. Because the study was partially unblinded for the three subjects dosed with NMD670 1600 mg or placebo, a new randomization for the remainder of their study visits was necessary. Therefore, the remaining two doses of Cohort 3 were randomized in a two-way cross-over design investigating 1200 mg NMD670 vs. placebo. Secondly, timing of PD measurements was updated for Cohort 3: the timing had been based on the expected time at which peak concentrations of NMD670 occurred (T_max_) based on animal data (around 1 hour). This was corrected to the actual timing based on the observed T_max_ in humans (3-4 hours).

### Study drug

Oral tablets of NMD670 50, 100 and 300 mg or matching placebo were administered with 240 ml water. Administered dose levels are shown in Figure S1. The study drug was administered as a single dose in the SAD study; once daily (QD) in the first three cohorts of the MAD study, and bidaily (BID) in the fourth cohort of the MAD study (the first and second dose were 6 hours apart). The starting dose of 50 mg was based on the NOAEL (pre-clinical no observed adverse effect level) approach and incorporated a 19-fold safety factor. The MABEL (minimal anticipated biological effect level) approach resulted in a similar starting dose of 49-97 mg. PAD (pharmacologically active dose) was estimated at 97-194 mg. The NMD670 fraction unbound (Fu) in vitro was 0.01 (high plasma protein binding), which was considered for the dose calculations. The randomization was generated in SAS (version 9.4) by a statistician uninvolved in study conduct. In the SAD study, there was a balanced assignment of three treatment sequences. Subjects and study team remained blinded during the study, with two exceptions made to assist with the selection of the next dose levels.

### Safety

Safety of NMD670 was investigated by monitoring adverse events (AE), vital signs, ECG, safety laboratory tests (blood chemistry, hematology, coagulation, urinalysis), and physical examinations. Moreover, due to the serum uric acid (sUA) decrease by NMD670 observed in SAD cohort 1 and 2, urinary uric acid and renal damage markers were measured in SAD Cohort 3 and MAD study. Additionally, urine was collected for 24 hours on the first and last MAD dosing day, to evaluate urinary pH and uric acid crystalloids. Later in the study, exploratory biomarkers (M35, M60, and GLDH) to evaluate drug-induced liver injury (DILI) were evaluated at Queen’s Medical Research, Edinburgh, UK.

As an exploratory safety measure, grip release profiles (GRREP) using handgrip dynamometry were used with the aim to detect subclinical myotonia ^5^. The measurement was performed using a handheld dynamometer (RS G200, Biometrics, Newport, UK) and wireless data transmitter (DataLite Pioneer – WS0, Biometrics). GRREP after three seconds of maximal voluntary contraction (MVC) was recorded (sampling rate 2000 Hz) and evaluated using in-house developed scripts. The MVC was defined as the median force applied during the last 500ms of the contraction period. GRREP was then characterized by the time it took to relax force production from 90%- MVC to 50%-MVC, and from 90%-MVC to 5%-MVC.

### Pharmacokinetics

The concentrations of NMD670 in human plasma (treated 1:1 (v/v) with Water: Orthophosphoric Acid (100:2), using EDTA as an anticoagulant) and urine (treated with 0.5 M Citric Acid: Tween-20 (95:5)), all samples were keep on ice and processed rapidly and stored frozen within validated time windows, was determined after solid-phase extraction followed by liquid chromatography tandem mass spectrometry (LC-MS/MS) by Labcorp Drug Development (Harrogate, UK) using a validated method. All plasma samples were analyzed within the validated storage period. The validated range for NMD670 was 50-50000 ng/mL and 300-150000 ng/mL in plasma and urine, respectively.

Non-compartmental analysis (NCA) of PK was performed using R (V4.0.3, R Core Team, Vienna, Austria) and the PKNCA package (v0.9.4) ^6^. Area under the concentration-time profiles (AUC) were derived using the linear-up log-down trapezoid rule. If half-life regression was successful, based on a minimum of 3 regression points, a R^2^ above 0.85 and a minimum span ratio of 1.5, the AUC_0-last_ was extrapolated to infinity to derive the AUC_inf_.

Dose proportionality of C_max_ and AUC_inf_/AUC_tau_ was evaluated graphically and quantitatively using a power model applied in SAS: LN(PK parameter)= a*LN(dose)+b, where a=slope and b=intercept. Dose proportionality was investigated by calculating if the 90% confidence interval (90%CI) of the slope of the regression was within the acceptance confidence interval defined as “1+*ln*(0-8)/*ln*(*dose*_*ratio*)<*slope*<1+*ln(1.25)/ln(dose_ratio)*” where dose_ratio is defined as the ratio between the highest and the lowest doses. To estimate a potential food effect, log-transformed AUC and C_max_ were compared with a mixed model analysis of variance, with food as fixed effect, and subject as random effect. To investigate a potential gender effect, log-transformed AUC and C_max_ were compared with a two-sample t-test between males and females at the same dose level (800 mg). To explore bioequivalence for food- and gender effects, the back-transformed 90%CI around the difference on log scale was calculated and compared with the 80.00% to 125.00% criteria.

Protein binding of NMD670 was determined using an HTDialysis 96-well plate consisting of teflon bars separated by individual cellulose dialysis membrane strips (12-14 kD molecular weight cut-off).

### Pharmacodynamics

Measurements of muscle velocity recovery cycles (MVRC) were performed to evaluate PD effects of NMD670 ^7^ ^8^. MVRC measurements were performed as described previously in the tibial muscle, using QTrac (recording protocol M3REC6, Institute of Neurology, London, UK) ^9^. Recovery cycles with 1, 2, and 5 conditioning stimuli (CS), as well as frequency ramp, were recorded. The following endpoints were generated for recovery cycles: relative refractory period (RRP); early supernormality after one (ESN) and five (5ESN) CS; interstimulus interval (ISI) corresponding to ESN (ESN@); supernormality at ISI 20 ms (SN20); late supernormality after one CS (LSN); difference in LSN after one versus two (XLSN) or five (5XLSN) CS; residual supernormality due to one CS (RSN); difference in RSN after one versus five CS (5XRSN). Endpoints for frequency ramp included: velocity change after stimulus trains of 15 Hz (Lat[15Hz]) and 30 Hz (Lat[30Hz]); action potential amplitude change after trains of 15 Hz (Peak[15Hz]) and 30 Hz (Peak[30Hz]); difference in amplitude 30 vs. 15 Hz (Pk[30-15Hz]); latency and amplitude change at 30 Hz vs 30 seconds after the ramp (Lat[30Hz30s] and Pk[30Hz30s]). The response to the first and last stimulus in the train is indicated with “_First_” and “_Last_”.

Before endpoints were generated, raw data was inspected by blinded staff. Abnormal muscle responses resulting in extreme outliers were interpolated. Additionally, a blinded data review was conducted on the endpoints to remove outliers in datapoints caused by technical issues.

Statistical analysis of PD endpoints was performed using a mixed effects model, with baseline as covariate, treatment, time, treatment by time as fixed factors, and subject as random factor. Variables were normally distributed.

## Results

A total of 35 subjects were included in the SAD study: 27 male, and eight female subjects. Moreover, 32 male subjects were included in the MAD study. Demographics are summarized in Table 1.

**Table 1:** Demographics of the single-ascending dose (SAD) study and multiple-ascending dose (MAD) study.

| Characteristic | Category/<br>Statistics | SAD study<br>N=35 | MAD study<br>N=32 |
| --- | --- | --- | --- |
| Sex | Female | 8 (22.9%) | 0 (0%) |
|  | Male | 27 (77.1%) | 32 (100%) |
| Race | Asian | 3 (8.6%) | 0 (0%) |
|  | Black | 1 (2.9%) | 1 (3.1%) |
|  | Mixed | 2 (5.7%) | 5 (15.6%) |
|  | Other | 1 (2.9%) | 0 (0%) |
|  | White | 28 (80.0%) | 26 (81.3%) |
| Age (y) | Mean (SD) | 34.7 (14.0) | 32.8 (15.2) |
|  | Range | 18, 63 | 19, 65 |
| Weight (kg) | Mean (SD) | 76.186 (10.249) | 75.130 (9.949) |
|  | Range | 58.00, 103.60 | 54.90, 98.30 |
| Height (cm) | Mean (SD) | 178.45 (9.79) | 180.12 (5.54) |
|  | Range | 155.5, 194.7 | 167.3, 192.5 |
| BMI (kg/m<sup>2</sup>) | Mean (SD) | 23.97 (3.05) | 23.15 (2.89) |
|  | Range | 19.4, 29.3 | 18.5, 30.0 |

**Abbreviations: BMI = body mass index; SD = standard deviation.**

### Safety

No serious AEs were reported in the SAD and the MAD studies.

*In the SAD study*, there were no meaningful relationships between increases in NMD670 dose and the incidence of participants with AEs (Table 2). A total of 70 AEs were reported, of which 47 (67%) were at least possibly drug related. The most commonly reported AEs (defined as those AEs reported in >1 subject) are listed in Table 2. There were no relationships between increases in dose and the incidence of these individual AEs following administration of a single dose of NMD670, except for transient myotonia which was reported at the highest dose levels tested (NMD670 1200 and 1600 mg). Most AEs were mild, except for one AE of myotonia (reported after 1600 mg NMD670), and tooth extraction (unrelated, reported after 50 mg NMD670), which were moderate in intensity. There were no severe AEs during the study and no subject discontinuations.

**Table 2:**
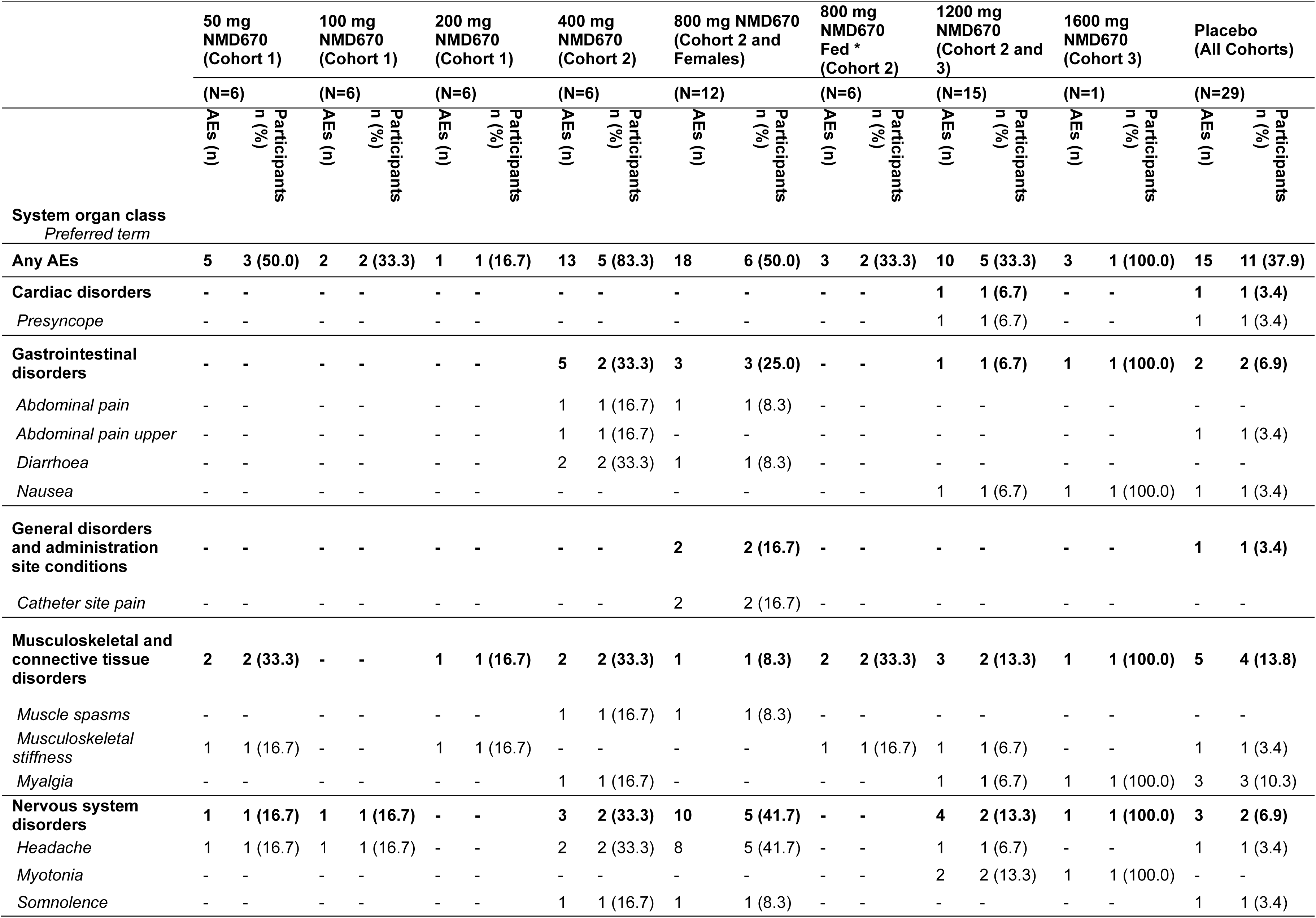
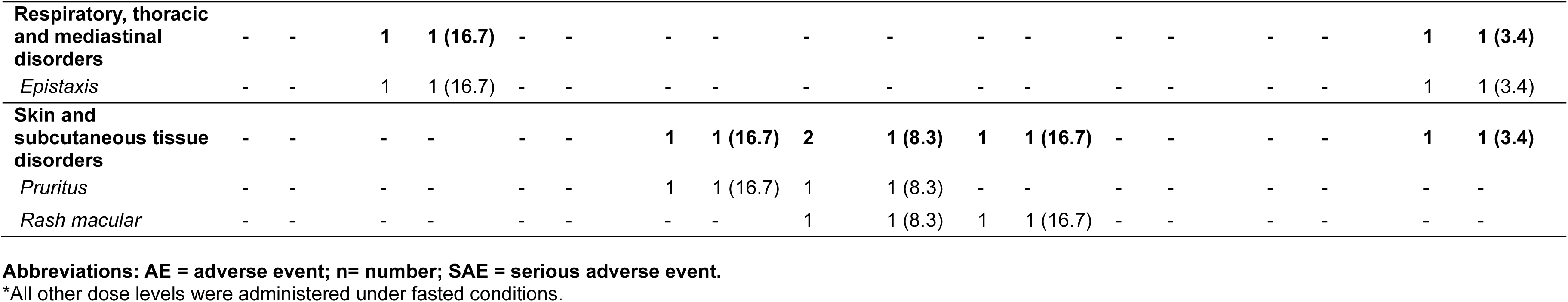
Most common adverse events (AEs) reported in the single-ascending dose study. This table shows AEs that were reported by more than one participant.

**Table 3:** Pharmacokinetic parameters of NMD670 in plasma and urine following administration of single ascending dose levels.

|  |  | 50 mg<br>NMD670<br>(Fasted) | 100 mg<br>NMD670<br>(Fasted) | 200 mg<br>NMD670<br>(Fasted) | 400 mg<br>NMD670<br>(Fasted) | 800 mg NMD670<br>(Fasted, Male) | 800 mg<br>NMD670<br>(Fed,<br>Male) | 800 mg NMD670<br>(Fasted, Female) | 1200 mg<br>(Fasted) | 1600 mg<br>(Fasted) |
| --- | --- | --- | --- | --- | --- | --- | --- | --- | --- | --- |
|  |  | N=6 | N=6 | N=6 | N=6 | N=6 | N=6 | N=6 | N=15 | N=1 |
| $C_{max}$<br>(ng/mL) | *Mean* | 1900.0 | 3978.3 | 7778.3 | 20050.0 | 36733.3 | 37266.7 | 44416.7 | 54980.0 | 123000.0 |
|  | *SD* | 579.90 | 1186.13 | 1128.72 | 7336.96 | 15362.90 | 7418.80 | 7477.28 | 21670.79 | - |
|  | *Min-max* | 1320-2890 | 2900-5940 | 5560-8600 | 10300-28100 | 15400-61800 | 29600-46500 | 34700-50900 | 23000-110000 | - |
| $T_{max}$ (h) | *Median* | 2.01 | 2.00 | 1.51 | 3.00 | 3.50 | 2.00 | 3.02 | 3.00 | 6.00 |
|  | *Min-max* | 1.00-4.00 | 1.00-4.00 | 1.00-4.00 | 1.00-4.00 | 3.00-4.08 | 1.00-6.00 | 2.00-4.02 | 1.00-6.00 | - |
| $AUC_{inf}$<br>(h*ng/mL) | *Mean* | - | 18487.8 | 36081.8 | 87727.2 | 190636.4 | 180132.3 | 255808.7 | 358473.6 | 826466.6 |
|  | *SD* | - | 1768.87 | 5033.86 | 21427.53 | 31524.38 | 27884.35 | 56186.23 | 176142.2 | - |
|  | *Min-max* | - | 16856-20930 | 31329-42851 | 66400-128033 | 160577-234933 | 140867-226164 | 183658-353759 | 208968-756825 | - |
| CL/F (L/h) | *Mean* | - | 5.445 | 5.626 | 4.753 | 4.289 | 4.530 | 3.249 | 3.896 | 1.936 |
|  | *SD* | - | 0.4964 | 0.7462 | 0.9723 | 0.6735 | 0.7009 | 0.6847 | 1.2783 | - |
|  | *Min-max* | - | 4.78-5.93 | 4.67-6.38 | 3.12-6.02 | 3.41-4.98 | 3.54-5.68 | 2.26-4.36 | 1.59-5.74 | - |
| $t_{1/2}$ (h) | *Mean* | - | 5.071 | 4.602 | 4.677 | 5.386 | 5.020 | 5.549 | 6.109 | 4.477 |
|  | *SD* | - | 0.6292 | 0.6710 | 0.4857 | 0.8152 | 1.0387 | 0.5511 | 1.2795 | - |
|  | *Min-max* | - | 4.43-5.73 | 3.93-5.73 | 4.08-5.37 | 4.23-6.42 | 3.91-6.32 | 4.57-6.03 | 3.44-8.39 | - |
| $V_z/F$ (L) | *Mean* | - | 39.73 | 37.54 | 32.08 | 33.09 | 32.25 | 25.66 | 34.53 | 12.50 |
|  | *SD* | - | 5.298 | 8.991 | 7.236 | 6.505 | 4.952 | 3.750 | 14.965 | - |
|  | *Min-max* | - | 34.5-47.0 | 30.6-52.7 | 20.6-39.1 | 29.1-46.1 | 25.4-39.7 | 19.0-29.6 | 11.3-69.2 | - |
| CLR (L/h) | *Mean* | 0.93 | 1.13 | 1.17 | 1.05 | 0.91 | - | - | 0.69 | 0.41 |
|  | *SD* | 0.37 | 0.48 | 0.44 | 0.60 | 0.67 | - | - | 0.36 | - |
|  | *Min-max* | 0.49-1.30 | 0.74-1.86 | 0.64-1.72 | 0.36-1.79 | 0.27-1.76 | - | - | 0.13-1.14 | - |

**Abbreviations:** $AUC_{inf}$ = area under the concentration-time curve from time zero to infinity; CL/F = apparent total clearance following extravascular administration; CLR = renal clearance; $C_{max}$ = maximum concentration; $V_z/F$ = apparent volume of distribution during the terminal elimination phase after extravascular administration. Concentrations below limit of quantification (25 ng/mL) were set to 0. For several dose levels, the N for certain parameters is smaller than the number listed at the top of the column.

*In the MAD study*, there were also no meaningful relationships between increases in dose and the incidence of participants with AEs (Table S1). A total of 65 AEs were reported, 37 (57%) were at least possibly drug related. Most common AEs (defined as those reported in >1 subject) were presyncope, catheter-site related reaction, fatigue, increased hepatic enzymes, musculoskeletal stiffness, myalgia, headache, and contact dermatitis. There were also no relationships between increases in dose and the incidence of these individual AEs following administration of multiple doses of NMD670. All AEs were mild in intensity. Two subjects discontinued the study: one subject on NMD670 discontinued due to an unrelated AE (influenza-like illness); one other subject on placebo withdrew consent.

Mild ALAT elevations up to two times upper limit of normal were observed in seven subjects throughout and after dosing, all returned to normal levels during the follow up period. In these subjects, bilirubin levels and all other liver function test (including coagulation times) remained normal during the study; exploratory DILI biomarkers (M65 and GLDH) showed possible mild liver injury. Of the affected subjects, two had received NMD670 200 mg QD, one received NMD670 600 mg QD, and four received placebo, thus none of the liver related safety abnormalities were considered compound related. There were no meaningful relationships between increasing dose and changes in the vital sign variables, nor ECG variables. No clinically significant abnormalities were reported in the Holter ECG recordings.

After a single dose of NMD670 1200 mg (N=15), mean sUA was reduced from 0.307 mmol/L pre-dose to 0.163 mmol/L 24 hours post-dose (mean change from baseline: 0.145 mmol/L; 47.2%), with a post-dose range of 0.10-0.25 mmol/L. Due to this observation, urinary uric acid determination was added from SAD Cohort 3 onwards, and increased uric acid excretion was observed. In the MAD study, reduction of sUA maintained throughout the ten dosing days, reaching a steady state after the first two to four days of dosing (Figure S2). Renal function markers such as urea, creatinine, cystatin C remained unremarkable across all treatment groups.

There was no overall effect of NMD670 on handgrip release times (Table S2). However, the 90%-MVC to 5%-MVC release time increased more than tenfold approximately six hours post-dose (corresponding to the personal T_max_) in the one participant who developed moderate symptoms of myotonia after NMD670 1600 mg administration. This was not observed in the two subjects with mild myotonia at 1200 mg.

### Pharmacokinetics

Summary PK profiles of all dose levels of NMD670 in the SAD and MAD study are shown in Figures 1 and 2, and PK parameters are shown in Tables 3 and S5. After a single dose of NMD670, the exposure increased in a dose dependent manner (Table S3). For AUC_inf_, the slope was 1.184 (90% CI: 1.090 to 1.277), which was fully outside the prespecified critical interval criteria (0.910 to 1.090). Results indicated a slightly more than dose proportional increase in AUCinf at higher doses. For C_max_, the slope of 1.050 (90% CI: 0.977 to 1.124) and the lower end of the CI were within the prespecified interval criteria (0.930 to 1.090), but the upper end was outside this interval, meaning dose proportionality could not be confirmed. The absorption of NMD670 had a median T_max_ of 1.5 to 3.5 hours. Plasma concentrations of NMD670 followed biexponential decline with mean apparent terminal half-life (T_1/2_) ranging between 4.6 and 6.1 hours. After multiple doses of NMD670, the exposure increased in a dose dependent manner. The absorption was similar to that found in the single dose study, with median T_max_ of NMD670 between 2.0 to 3.5 hours.

**Figure 1:**
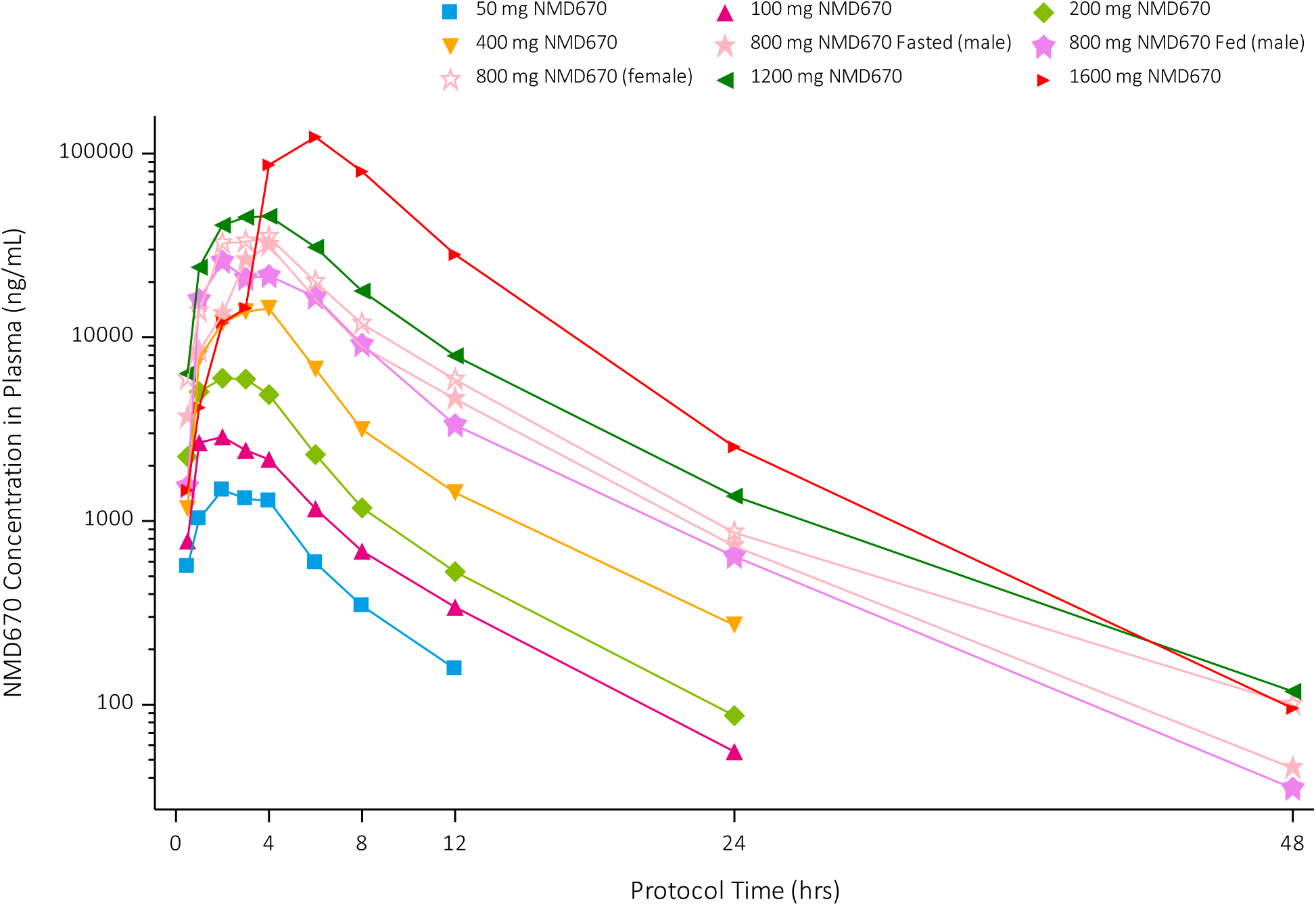
Mean of NMD670 concentrations in plasma (ng/mL) for all single-ascending dose levels, presented on a semi-log scale.

**Figure 2:**
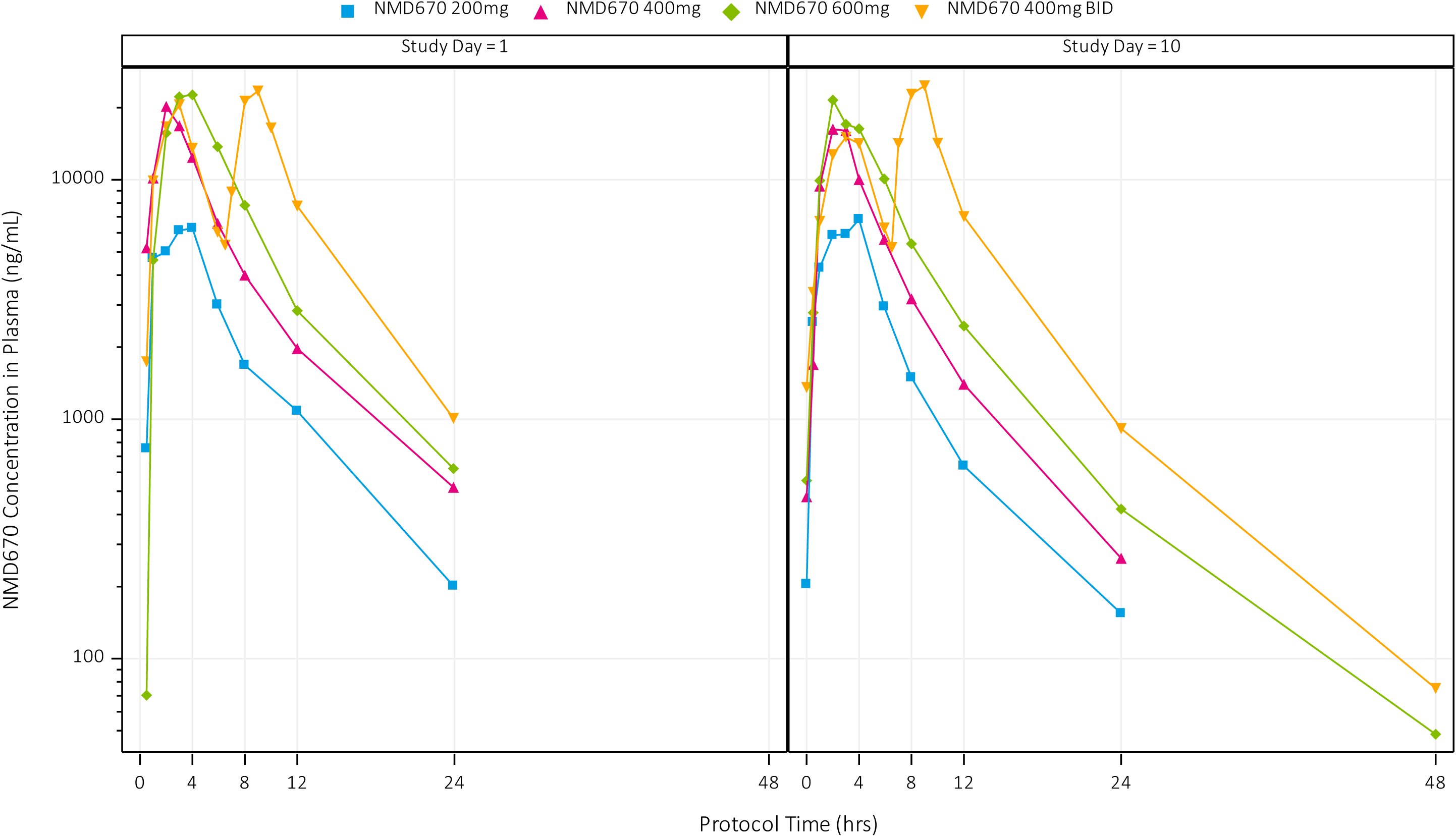
Mean of NMD670 concentrations in plasma (ng/mL) for all multiple-ascending dose levels, presented on a semi-log scale on Day 1 (first day of dosing) and Day 10 (last day of dosing).

Steady state (for QD dosing) was reached after the first dosing for NMD670. No accumulation of NMD670 was observed between single and multiple dosing, with a mean Rac(AUC) ranging from 0.798 to 0.955 for QD and BID dosing; the mean Rac(C_max_) ranged from 0.811 to 1.113 for QD and BID dosing.

As judged by the AUC_inf_ ratio, the extent of absorption was not influenced by food (Table S4). The variability in C_max_ resulted in a broad 90% CI, so the presence of a food effect could not be confirmed for this parameter. Moreover, our results show a significant difference in AUC_inf_ between genders, with a higher exposure in females. For C_max_, a gender effect could not be confirmed as the upper end of the CI was outside the interval criteria. Of note, there were differences in demographics between male and female subjects: mean age, weight, and BMI was for males 28 years, 77.0 kg, and 23.2 kg/m2, respectively; and for females 56 years, 73.4 kg, and 26.6 kg/m2, respectively.

Mean percentage of administered dose excreted into the urine from time zero to time of last measurable concentration (Ae%last) was 19.0% (Cohort 3). Mean fraction unbound (Fu) was 1.14 ± 0.46% in male, and 1.18 ± 0.16% in female participants. Mean observed renal clearance was between 0.69 ± 0.36 and 1.17 ± 0.44 L/h. This is higher than the expected renal clearance if only dependent on passive filtration, calculated as the glomerular filtration rate (assumed to be 125 mL/min) times Fu, suggesting active secretion of NMD670 in urine.

The two subjects who developed mild myotonia at NMD670 1200 mg had a higher-than-average C_max_: 85,200 and 110,000 ng/mL (average C_max_ for 1200 mg was 54,980 ng/mL).

### Pharmacodynamics

No consistent significant effect of NMD670 in increasing dose levels was observed on MVRC variables in SAD Cohort 1 and 2 (Table S6). This in contrast to Cohort 3 (two-way crossover), where significant effects of NMD670 1200 mg vs placebo were observed (Table 4). The recovery cycles showed a significant increase in ESN (*p*=0.024) and 5ESN (*p*=0.018) with NMD670 compared to placebo. Moreover, SN20 was also significantly increased after NMD670 administration compared with placebo (*p*=0.002). The frequency ramp showed significantly decreased Lat[15Hz]last (*p*=0.034) and increased Lat[30Hz+30s] (*p*=0.003) after NMD670. Average MVRC recordings in Figure 3 visualize these effects.

**Figure 3:**
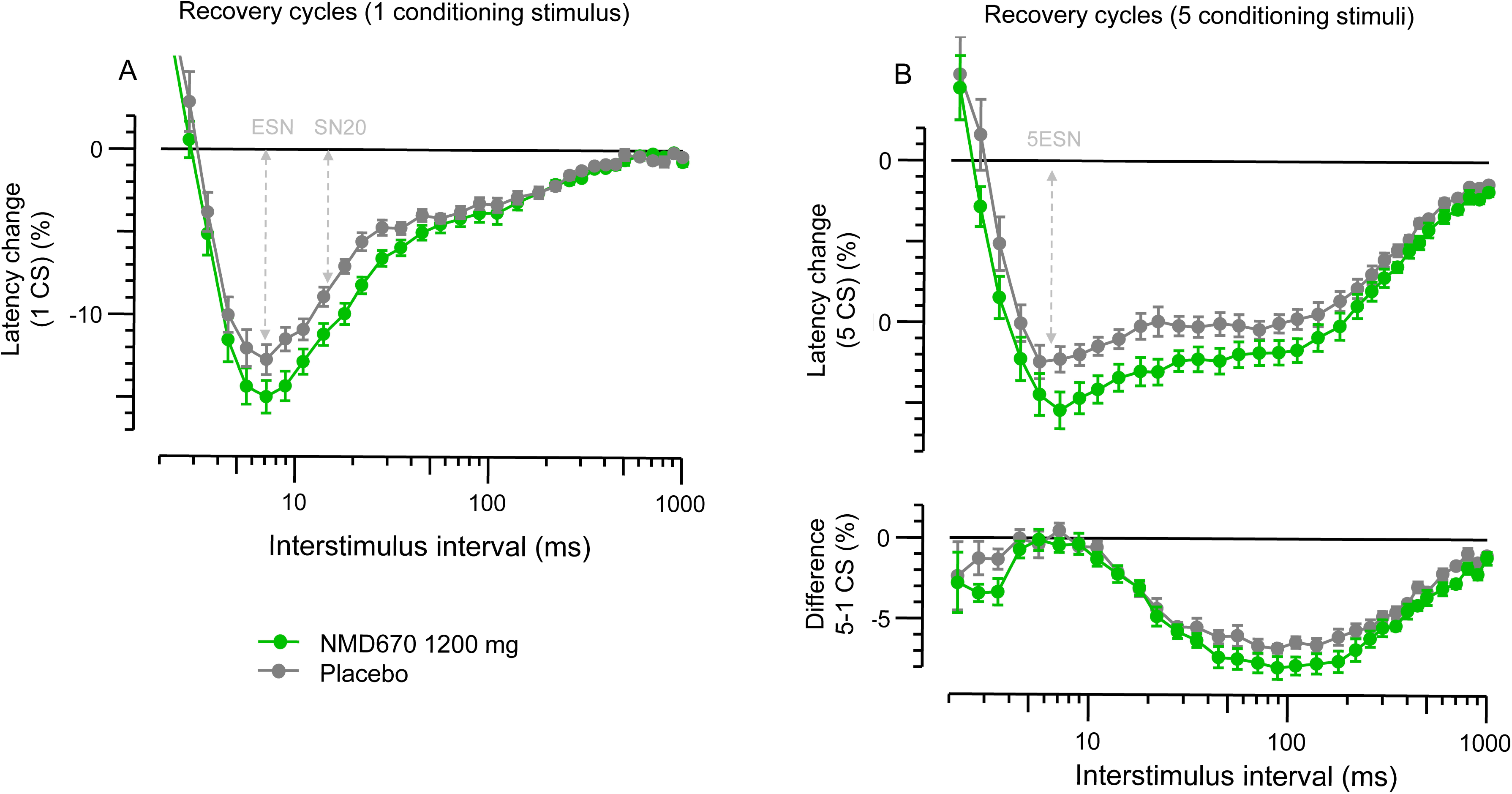
Mean recordings of recovery cycles, as measured post-dose for NMD670 1200 mg (green) and placebo (grey). A) Percentual latency change after one conditioning stimulus at different interstimulus intervals. B) Upper graph: the percentual latency change after five conditioning stimuli at different interstimulus intervals; Lower graph: the difference in latency change, between five and one conditioning stimuli. The standard error is visualized in error bars, statistically significant findings of NMD670 vs. placebo are highlighted. This graph does not reflect the statistical analysis, because the statistical model includes baseline as a covariate. Visualization of parameters reproduced from Tan *et al*., 2014.

**Table 4:**
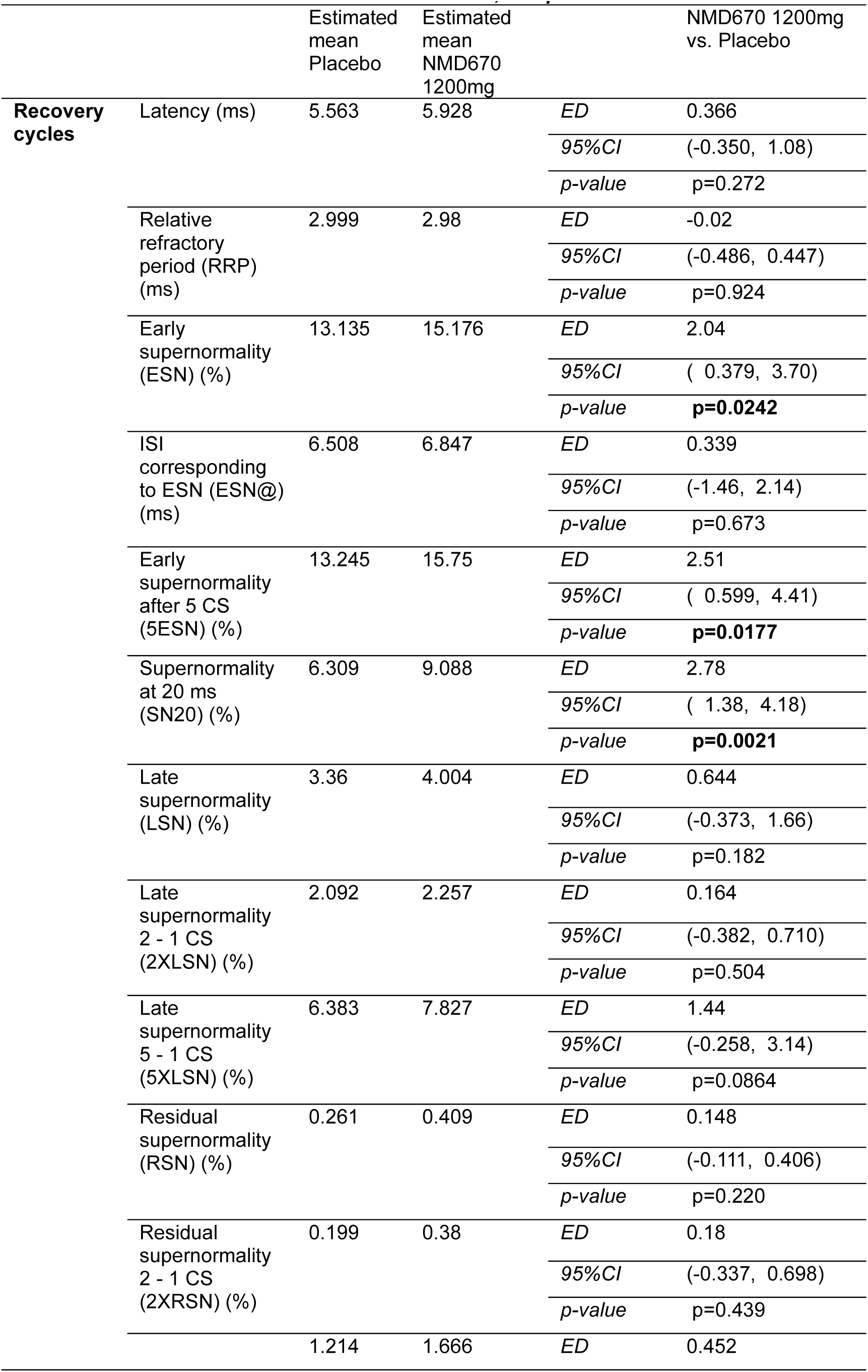
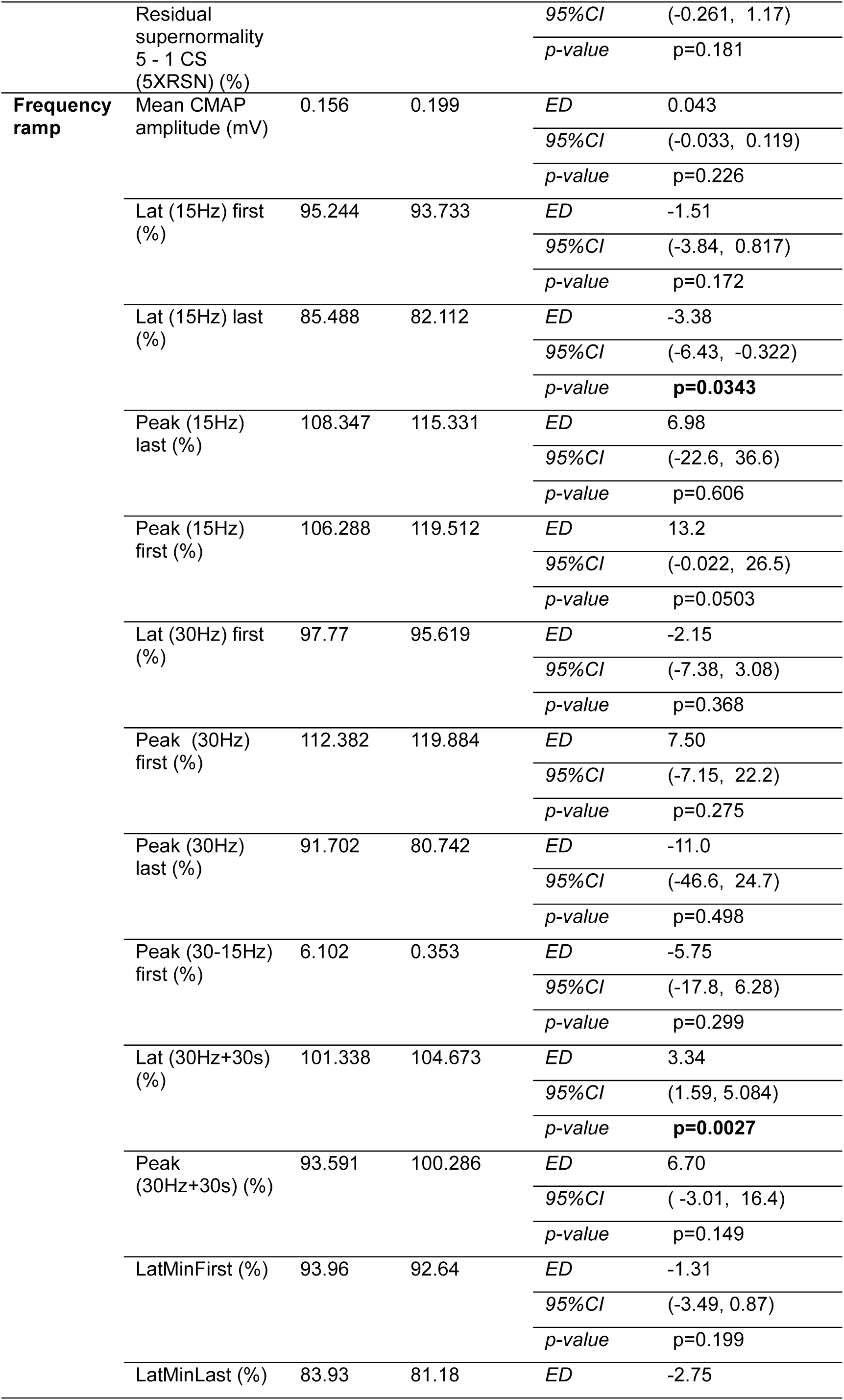
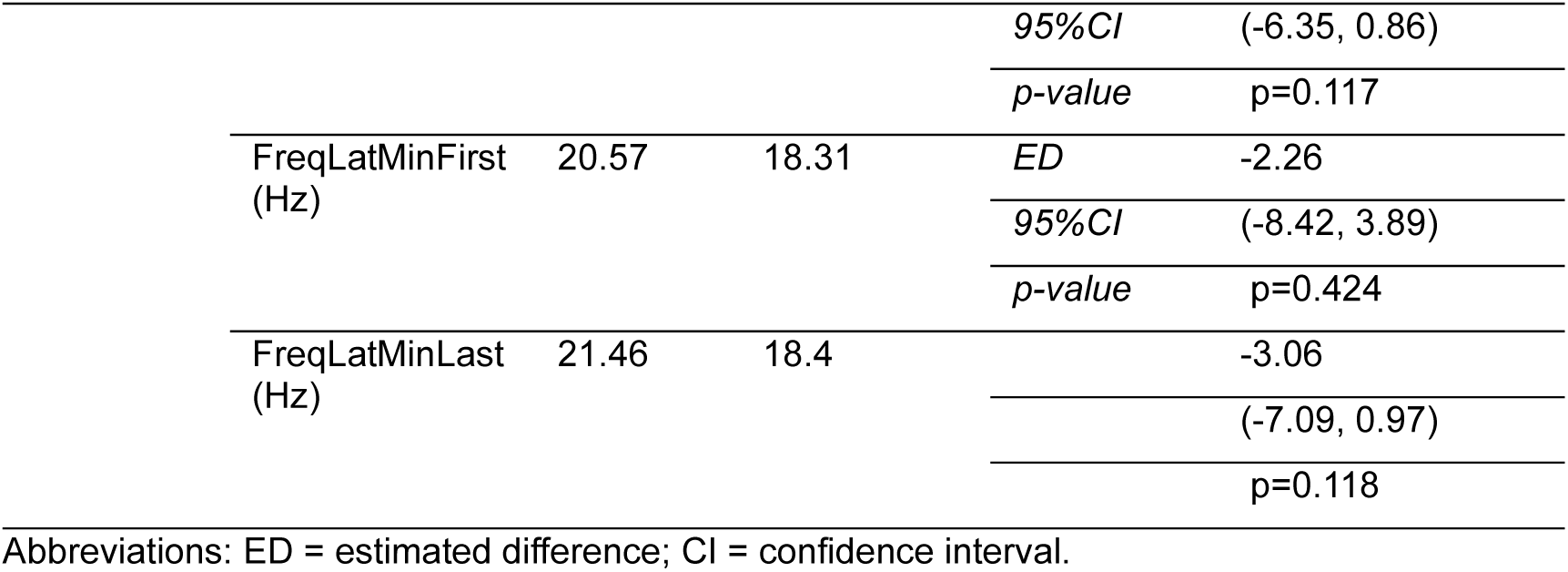
Effects of a single dose of NMD670 1200 mg (Cohort 3) versus placebo on muscle velocity recovery cycles. The table shows estimated means post-dose for placebo and NMD670; and the estimated difference, with 95% confidence interval of the estimated mean, and *p*-value.

|  |  | Estimated mean<br>Placebo | Estimated mean<br>NMD670<br>1200mg |  | NMD670 1200mg<br>vs. Placebo |
| --- | --- | --- | --- | --- | --- |
| **Recovery**<br>**cycles** | Latency (ms) | 5.563 | 5.928 | *ED* | 0.366 |
|  |  |  |  | *95%CI* | (-0.350, 1.08) |
|  |  |  |  | *p-value* | p=0.272 |
|  | Relative refractory period (RRP) (ms) | 2.999 | 2.98 | *ED* | -0.02 |
|  |  |  |  | *95%CI* | (-0.486, 0.447) |
|  |  |  |  | *p-value* | p=0.924 |
|  | Early supernormality (ESN) (%) | 13.135 | 15.176 | *ED* | 2.04 |
|  |  |  |  | *95%CI* | ( 0.379, 3.70) |
|  |  |  |  | *p-value* | **p=0.0242** |
|  | ISI corresponding to ESN (ESN@) (ms) | 6.508 | 6.847 | *ED* | 0.339 |
|  |  |  |  | *95%CI* | (-1.46, 2.14) |
|  |  |  |  | *p-value* | p=0.673 |
|  | Early supernormality after 5 CS (5ESN) (%) | 13.245 | 15.75 | *ED* | 2.51 |
|  |  |  |  | *95%CI* | ( 0.599, 4.41) |
|  |  |  |  | *p-value* | **p=0.0177** |
|  | Supernormality at 20 ms (SN20) (%) | 6.309 | 9.088 | *ED* | 2.78 |
|  |  |  |  | *95%CI* | ( 1.38, 4.18) |
|  |  |  |  | *p-value* | **p=0.0021** |
|  | Late supernormality (LSN) (%) | 3.36 | 4.004 | *ED* | 0.644 |
|  |  |  |  | *95%CI* | (-0.373, 1.66) |
|  |  |  |  | *p-value* | p=0.182 |
|  | Late supernormality 2 - 1 CS (2XLSN) (%) | 2.092 | 2.257 | *ED* | 0.164 |
|  |  |  |  | *95%CI* | (-0.382, 0.710) |
|  |  |  |  | *p-value* | p=0.504 |
|  | Late supernormality 5 - 1 CS (5XLSN) (%) | 6.383 | 7.827 | *ED* | 1.44 |
|  |  |  |  | *95%CI* | (-0.258, 3.14) |
|  |  |  |  | *p-value* | p=0.0864 |
|  | Residual supernormality (RSN) (%) | 0.261 | 0.409 | *ED* | 0.148 |
|  |  |  |  | *95%CI* | (-0.111, 0.406) |
|  |  |  |  | *p-value* | p=0.220 |
|  | Residual supernormality 2 - 1 CS (2XRSN) (%) | 0.199 | 0.38 | *ED* | 0.18 |
|  |  |  |  | *95%CI* | (-0.337, 0.698) |
|  |  |  |  | *p-value* | p=0.439 |
|  |  | 1.214 | 1.666 | *ED* | 0.452 |
|  | Residual<br>supernormality<br>5 - 1 CS<br>(5XRSN) (%) |  |  | 95%CI | (-0.261, 1.17) |
|  |  |  |  | p-value | p=0.181 |
| **Frequency**<br>**ramp** | Mean CMAP<br>amplitude (mV) | 0.156 | 0.199 | ED | 0.043 |
|  |  |  |  | 95%CI | (-0.033, 0.119) |
|  |  |  |  | p-value | p=0.226 |
|  | Lat (15Hz) first<br>(%) | 95.244 | 93.733 | ED | -1.51 |
|  |  |  |  | 95%CI | (-3.84, 0.817) |
|  |  |  |  | p-value | p=0.172 |
|  | Lat (15Hz) last<br>(%) | 85.488 | 82.112 | ED | -3.38 |
|  |  |  |  | 95%CI | (-6.43, -0.322) |
|  |  |  |  | p-value | **p=0.0343** |
|  | Peak (15Hz)<br>last (%) | 108.347 | 115.331 | ED | 6.98 |
|  |  |  |  | 95%CI | (-22.6, 36.6) |
|  |  |  |  | p-value | p=0.606 |
|  | Peak (15Hz)<br>first (%) | 106.288 | 119.512 | ED | 13.2 |
|  |  |  |  | 95%CI | (-0.022, 26.5) |
|  |  |  |  | p-value | p=0.0503 |
|  | Lat (30Hz) first<br>(%) | 97.77 | 95.619 | ED | -2.15 |
|  |  |  |  | 95%CI | (-7.38, 3.08) |
|  |  |  |  | p-value | p=0.368 |
|  | Peak (30Hz)<br>first (%) | 112.382 | 119.884 | ED | 7.50 |
|  |  |  |  | 95%CI | (-7.15, 22.2) |
|  |  |  |  | p-value | p=0.275 |
|  | Peak (30Hz)<br>last (%) | 91.702 | 80.742 | ED | -11.0 |
|  |  |  |  | 95%CI | (-46.6, 24.7) |
|  |  |  |  | p-value | p=0.498 |
|  | Peak (30-15Hz)<br>first (%) | 6.102 | 0.353 | ED | -5.75 |
|  |  |  |  | 95%CI | (-17.8, 6.28) |
|  |  |  |  | p-value | p=0.299 |
|  | Lat (30Hz+30s)<br>(%) | 101.338 | 104.673 | ED | 3.34 |
|  |  |  |  | 95%CI | (1.59, 5.084) |
|  |  |  |  | p-value | **p=0.0027** |
|  | Peak<br>(30Hz+30s) (%) | 93.591 | 100.286 | ED | 6.70 |
|  |  |  |  | 95%CI | ( -3.01, 16.4) |
|  |  |  |  | p-value | p=0.149 |
|  | LatMinFirst (%) | 93.96 | 92.64 | ED | -1.31 |
|  |  |  |  | 95%CI | (-3.49, 0.87) |
|  |  |  |  | p-value | p=0.199 |
|  | LatMinLast (%) | 83.93 | 81.18 | ED | -2.75 |

|  |  |  |  |  |
| --- | --- | --- | --- | --- |
|  |  |  | *95%CI* | (-6.35, 0.86) |
|  |  |  | *p-value* | p=0.117 |
| FreqLatMinFirst<br>(Hz) | 20.57 | 18.31 | *ED* | -2.26 |
|  |  |  | *95%CI* | (-8.42, 3.89) |
|  |  |  | *p-value* | p=0.424 |
| FreqLatMinLast<br>(Hz) | 21.46 | 18.4 |  | -3.06 |
|  |  |  |  | (-7.09, 0.97) |
|  |  |  |  | p=0.118 |

Abbreviations: ED = estimated difference; CI = confidence interval.

There were no systematic treatment effects on MVRC parameters at any dose in the MAD study (Table S6).

## Discussion

NMD670, an inhibitor of ClC-1, is a novel first-in-class compound in development for symptomatic treatment of NMDs such as MG. We describe the safety, PK, and PD of NMD670 in healthy subjects.

NMD670 was generally safe and well-tolerated in healthy subjects, and no relation of AEs with dose was observed, with the exception of myotonia occurring at the highest dose levels tested. Myotonia resolved fully and spontaneously within hours, and it was not considered a safety hazard to the subject. NMD670 1200 mg was set as the maximum tolerated dose in healthy subjects due to the incidence of moderate myotonia at NMD670 1600 mg.

Importantly, the symptoms of myotonia indicate target engagement of NMD670 as ClC-1 inhibitor. In fact, impaired function of ClC-1 due to loss-of-function mutations, is known to cause myotonia congenita, because of increased muscle cell excitability ^10^. It should be noted that previous reports have demonstrated that symptoms of myotonia appear when more than 80% of the ClC-1 function has been compromised ^1^. The observed effect of NMD670 at the highest dosing in the present study would therefore suggest a substantial level of ClC-1 inhibition, considerably exceeding the level of block required to demonstrate recovery of neuromuscular transmission in pre-clinical models of MG, which is in the range of 20% inhibition. ^11,12^ The observation of increased handgrip release times in the participant with myotonia after 1600 mg NMD670 further indicates intended on-target pharmacology.

NMD670 (1200 mg) induced significant effects on MVRC variables. Previous work shows that MVRC can be used to detect acute pharmacological effects of drugs that modify muscle excitability ^9^. The MVRC method was expected to be sensitive to inhibition of ClC-1 because effects of decreased ClC-1 function in patients with myotonia congenita can be demonstrated using MVRC ^8^. Our study showed that NMD670 increased MVRC variables ESN, 5ESN and SN20, indicating increased muscle cell excitability. In myotonia congenita (compared to healthy controls) ESN, 5ESN and SN20 were also increased, and Lat[15Hz]_last_ decreased ^8^ , strengthening the hypothesis that the effects observed with NMD670 are indeed a result of ClC-1 inhibition. Together, these results provide further evidence of pharmacological target engagement in healthy subjects at safe and tolerable dose levels.

No consistent or dose-dependent effects of NMD670 on MVRC parameters were detected in SAD Cohorts 1 and 2, or in the MAD. Levels of ClC-1 inhibition in SAD Cohorts 1 and 2 and the MAD study may have been insufficient to induce detectable effects with MVRC, because the dose levels were lower than the dose level (1200 mg) where significant PD effects were observed in the SAD study. However, it is also important to note that MVRC measurements in SAD Cohort 1 and 2 were performed earlier than T_max_ (1-hour post-dose), and the timing was adjusted to the observed PK in Cohort 3 to coincide with T_max_ (3 hours post-dose), thereby better aligning the PD with the optimal PK. Moreover, as the MAD employed a parallel design, it had substantially less power to detect effects than the cross-over design in SAD Cohort 3. Therefore, concentration-dependent effects on MVRC may have been present, but the design of SAD Cohort 1 and 2 and the MAD did not allow to test this hypothesis adequately.

Another possible limitation of the PD analysis is the lack of multiple testing corrections, generally accepted in Phase I studies due to the exploratory nature and small sample size of the study.

The observed PK profile and the dose dependent increase in exposure allows for QD and BID dosing in a wide range of exposure levels in future studies. Investigation of food and gender effects on PK was exploratory. With sample size n=6, the study was not fully powered to assess bioequivalence. Even though a significant difference in AUC_inf_ ratio between male and female subjects was observed, this could be driven by a difference in age and BMI between the two groups.

NMD670 led to a dose-dependent increase in uric acid excretion. The likely mechanism of this change is inhibition of anion-exchanging uptake transporter URAT1 in the proximal tubular cells by NMD670, a transporter responsible for most of the uric acid reabsorption ^13^. Whether increased uric acid excretion is a safety concern is unclear. Hereditary renal hypouricemia is associated with exercise-induced acute kidney injury and kidney stones ^14^ and hypouricemia has been associated with higher mortality ^15^ and progression of neurodegenerative disorders ^16^, but the evidence is limited and inconsistent. Our data shows no indication of complications due to low sUA, neither in the acute renal excretion phase, nor during ten days of dosing. Future studies with NMD670 will be needed to evaluate effects of long-term use.

Lastly, mild and transient elevations of ALAT and M65 and GLDH changes (exploratory biomarkers of liver function) were observed after multiple dosing. As these occurred with a higher absolute and relative frequency in the placebo group, these were deemed to be very unlikely related to NMD670 administration.

### Conclusion

This investigation describes the first single- and multiple dose administration of a first-in-class selective ClC-1 channel inhibitor (NMD670) in healthy subjects. Inhibition of ClC-1 may improve the transmission of action potentials in the neuromuscular junction through increased muscle excitability, making ClC-1 an interesting novel target for the treatment of NMDs, including but not limited to MG. NMD670 was generally safe and well-tolerated, with expected mild to moderate myotonia at the higher dose levels, reflecting exaggerated on-target pharmacology, which resolved spontaneously within hours. Moreover, NMD670 showed significant effects on MVRC parameters, namely increased ESN, 5ESN and SN20, indicating pharmacological target engagement. The current study in healthy subjects provides a solid base for translation to patients, with proof-of-pharmacology and evidence of target engagement at dose levels that were considered safe and well-tolerated. Further investigations in patients with neuromuscular disorders are warranted and should confirm whether the mechanism of ClC-1 inhibition improves neuromuscular transmission deficits in association with increases of muscle strength and function in these devastating disorders.

## Study highlights

### What is the current knowledge on the topic?

There is a clear unmet need for tolerable treatments for neuromuscular diseases, such as myasthenia gravis. Animal studies show that inhibition of ClC-1 enhances muscle excitability, which improves transmission of action potentials at the neuromuscular junction and, consequently, muscle function. NMD670 is a first-in-class selective ClC-1 inhibitor developed to improve muscle weakness and fatigue.

### What question did this study address?

The safety, pharmacokinetics, and pharmacodynamics of single and multiple doses of NMD670 were evaluated in healthy male and female participants.

### What does this study add to our knowledge?

This paper describes the first single- and multiple dose administration of NMD670 in healthy subjects. NMD670 was generally safe and well-tolerated. NMD670 caused mild to moderate myotonia at the highest dose levels and showed target engagement on MVRC.

### How might this change clinical pharmacology or translational science?

The current study in healthy subjects provides a solid base for translation to patients, with evidence of target engagement at dose levels that were considered safe and well-tolerated in healthy subjects. Patient studies should confirm whether the mechanism of ClC-1 inhibition improves neuromuscular transmission deficits and provides clinical benefits.

## Data Availability

All data needed to interprete the work are contained in the manuscript

## Conflict of interest

This study was sponsored by NMD Pharma A/S. The funder had the following involvement with the study: study design, study oversight and medical monitoring, representation in dose escalation committee, decision to publish, and preparation of the manuscript. When this work was performed, JH, JB, TSG, KG, EC, JQ, PF, and TP were consultants or full-time employees of NMD Pharma who may own and/or hold options/restricted stock units for the company. All other authors declared no competing interests for this work.

## Funding information

The study was sponsored by NMD Pharma A/S. Prof. G.J. Groeneveld was Principle Investigator of this study.

## Author contributions

T.R., C.C., J.A.H., G.J.G. wrote the first draft of the manuscript. T.R., J.A.H., J.H., J.B., T.G., K.J., E.C., R.J.D., T.H.P. and G.J.G. designed the research. T.R., C.C., I.K., A.G., and G.J.G. performed the research. M.K., M.E., E.K. analyzed the data. All authors critically reviewed the manuscript and approved the final version for publication.

## Data accessibility statement

The study protocol, and data and scripts that support the findings of this study, are available from the corresponding author upon reasonable request.

## Supplementary material

Figure S1: Schematic overview of the study design. The single-ascending dose study followed a partial cross-over randomization. Dose levels highlighted in grey indicate a different set-up, namely: 1) Cohort 2, in which subjects that had received 800 mg in a fasted state, received NMD670 800 mg in fed state in the same randomization ; 2) the last two doses of Cohort 3, in which subjects received 1200 mg and placebo in a full cross-over randomization; 3) the female cohort in which subjects received a single dose of NMD670 or placebo (randomization 6:2 for NMD670 vs. placebo). Subjects in the multiple-ascending dose study were randomized in a parallel study design. Abbreviations: BID = bis in diem/twice a day; QD = quaque die/once a day.

Figure S2: Change from baseline (CFB) serum uric acid levels (mmol/L) following multiple-dose administrations of NMD670 or placebo.

Table S1: Most common adverse events reported in the multiple ascending dose study. This table shows AEs that were reported by more than one participant.

Table S2: Effects of NMD670 versus placebo on handgrip release profile, for single- and multiple-ascending doses. The table shows post-dose estimated means for placebo and NMD670; and the estimated difference, with 95% confidence interval of the estimated mean, and p-value.

Table S3: Evaluation of dose proportionality for pharmacokinetic parameters AUCinf and C_max_. Dose proportionality can be confirmed when the 90% confidence interval (90% CI) of the slope of the regression falls within the critical confidence interval (CI).

Table S4: Evaluation of food and gender effects on pharmacokinetic parameters AUCinf and C_max_. To explore bioequivalence the back-transformed 90% confidence interval (CI) around the difference on log scale was compared with the 80.00% to 125.00% criteria.

Table S5: Pharmacokinetic parameters of NMD670 in plasma following administration of all multiple ascending dose levels.

Table S6 Effects of NMD670 versus placebo on muscle velocity recovery cycles, in single- and multiple-ascending doses. The table shows post-dose estimated means for placebo and NMD670; and the estimated difference, with 95% confidence interval of the estimated mean, and p-value.

## Notes

### Clinical Trial

NL8692

### Funding Statement

The study was funded by NMD Pharma A/S

### Author Declarations

Ethics Committee Stichting of Beoordeling Ethiek Biomedisch Onderzoek The Netherlands gave ethical approval for this work

