## Supplementary Material for "SAFETY, PHARMACOKINETICS, AND PHARMACODYNAMICS OF A CLC-1 INHIBITOR - A FIRST-IN-CLASS COMPOUND THAT ENHANCES MUSCLE EXCITABILITY: A PHASE I, SINGLE- AND MULTIPLE-ASCENDING DOSE STUDY"

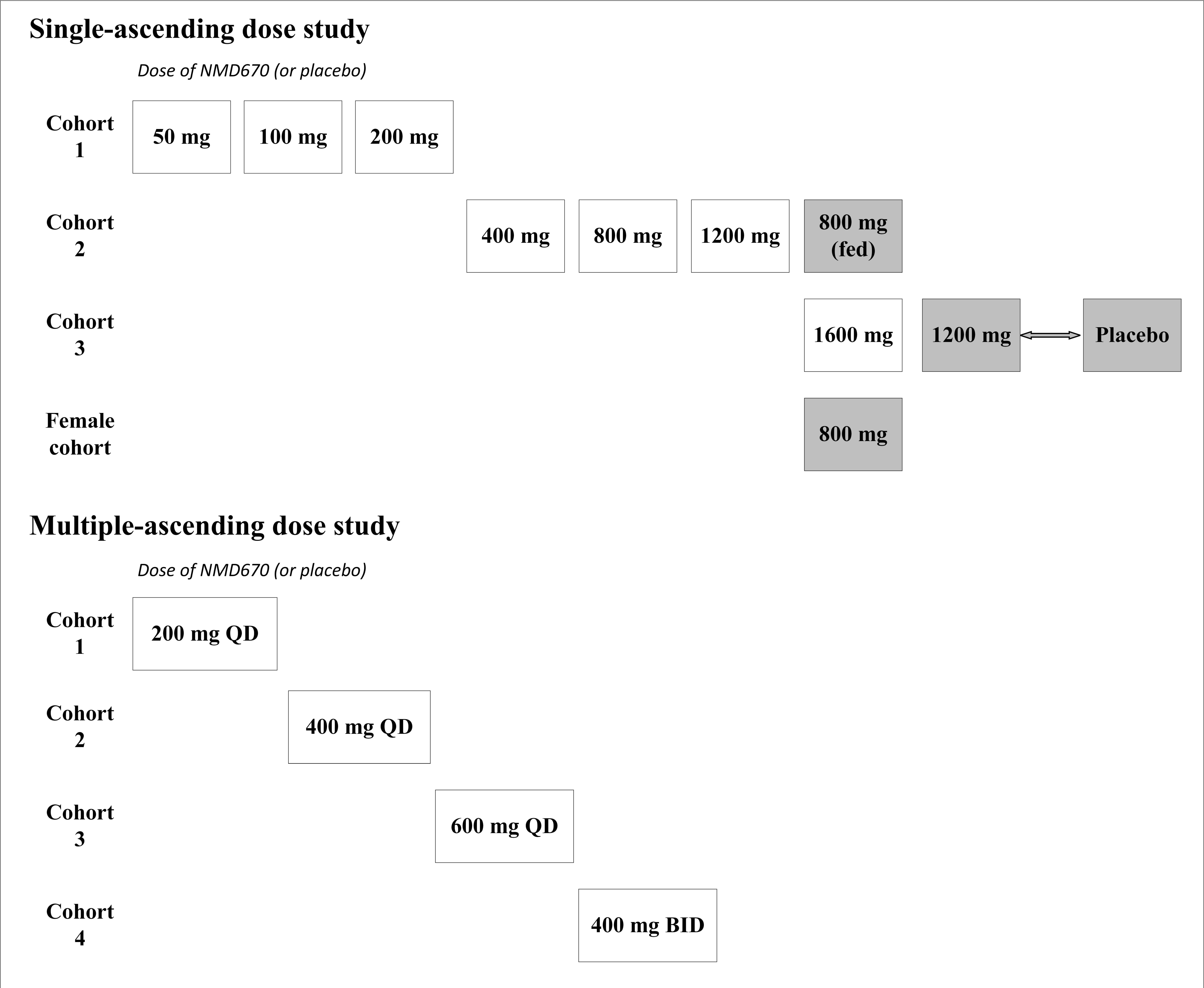
Single-ascending dose study
Dose of NMD670 (or placebo)
Cohort 1
50 mg
100 mg
200 mg
Cohort 2
400 mg
800 mg
1200 mg
800 mg (fed)
Cohort 3
1600 mg
1200 mg
Placebo
Female cohort
800 mg
Multiple-ascending dose study
Dose of NMD670 (or placebo)
Cohort 1
200 mg QD
Cohort 2
400 mg QD
Cohort 3
600 mg QD
Cohort 4
400 mg BID

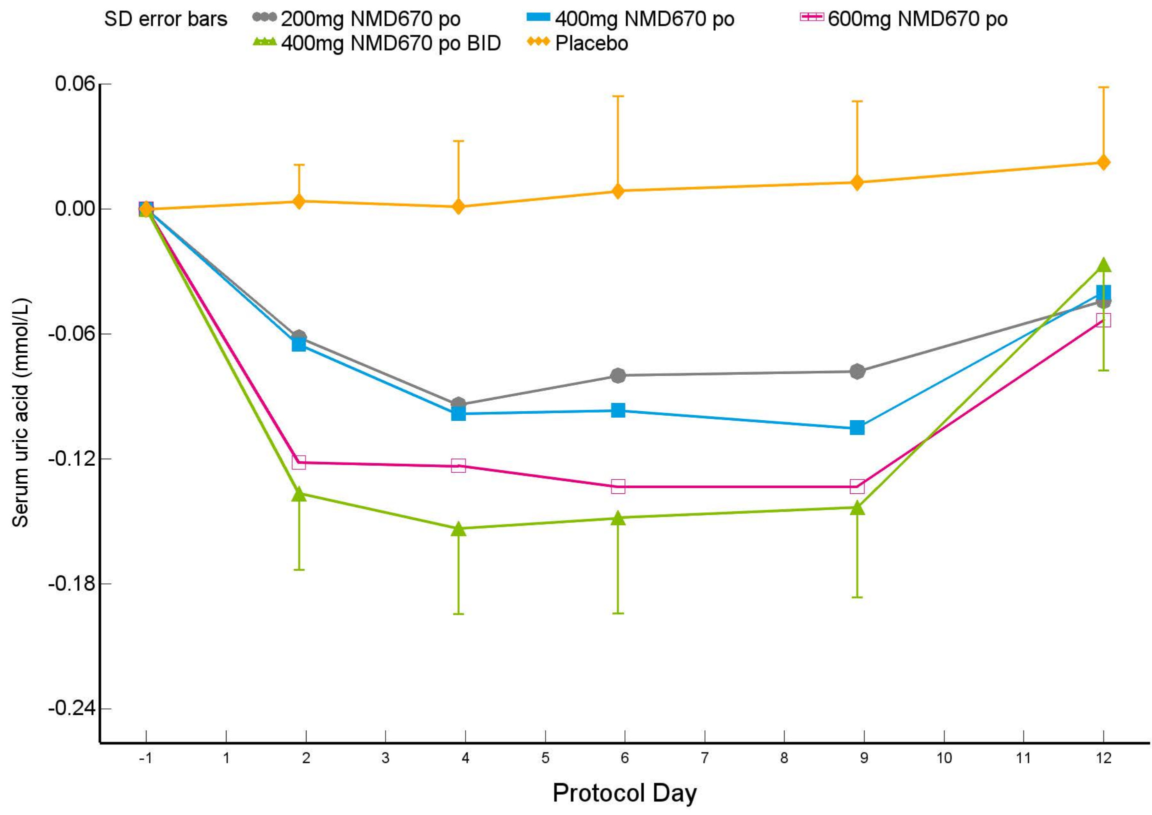
SD error bars
200mg NMD670 po
400mg NMD670 po
600mg NMD670 po
400mg NMD670 po BID
Placebo
0.06
0.00
-0.06
-0.12
-0.18
-0.24
Serum uric acid (mmol/L)
-1
1
2
3
4
5
6
7
8
9
10
11
12
Protocol Day

**Table S1: Most common adverse events reported in the multiple ascending dose study. This table shows AEs that were reported by more than one participant.**

| | **200 mg NMD670 QD (Cohort 1)** | | **400 mg NMD670 QD (Cohort 2)** | | **600 mg NMD670 QD (Cohort 3)** | | **400 mg NMD670 BID (Cohort 4)** | | **Placebo (All Cohorts)** | |
|---|---|---|---|---|---|---|---|---|---|---|
| | **(N=6)** | | **(N=6)** | | **(N=6)** | | **(N=6)** | | **(N=8)** | |
| **System organ class**<br>*Preferred term* | **AEs (n)** | **Participants (n [%])** | **AEs (n)** | **Participants (n [%])** | **AEs (n)** | **Participants (n [%])** | **AEs (n)** | **Participants (n [%])** | **AEs (n)** | **Participants (n [%])** |
| **Any events** | **13** | **6 (100.0)** | **5** | **3 (50.0)** | **12** | **5 (83.3)** | **10** | **3 (50.0)** | **25** | **8 (100.0)** |
| **Cardiac disorders** | **1** | **1 (16.7)** | **1** | **1 (16.7)** | **-** | **-** | **-** | **-** | **-** | **-** |
| *Presyncope* | 1 | 1 (16.7) | 1 | 1 (16.7) | - | - | - | - | - | - |
| **General disorders and administration site conditions** | **2** | **2 (33.3)** | **-** | **-** | **4** | **2 (33.3)** | **-** | **-** | **6** | **5 (62.5)** |
| *Catheter site related reaction* | - | - | - | - | 2 | 1 (16.7) | - | - | - | - |
| *Fatigue* | - | - | - | - | 1 | 1 (16.7) | - | - | 3 | 3 (37.5) |
| **Investigations** | **2** | **2 (33.3)** | **-** | **-** | **1** | **1 (16.7)** | **-** | **-** | **5** | **4 (50.0)** |
| *Hepatic enzyme increased* | 1 | 1 (16.7) | - | - | 1 | 1 (16.7) | - | - | 4 | 4 (50.0) |
| **Musculoskeletal and connective tissue disorders** | **-** | **-** | **3** | **3 (50.0)** | **-** | **-** | **3** | **2 (33.3)** | **3** | **3 (37.5)** |
| *Musculoskeletal stiffness* | - | - | 1 | 1 (16.7) | - | - | - | - | 1 | 1 (12.5) |
| *Myalgia* | - | - | 1 | 1 (16.7) | - | - | 1 | 1 (16.7) | - | - |
| **Nervous system disorders** | 4 | 2 (33.3) | 1 | 1 (16.7) | 3 | 3 (50.0) | 2 | 1 (16.7) | 5 | 4 (50.0) |
| *Headache* | 4 | 2 (33.3) | - | - | 3 | 3 (50.0) | 2 | 1 (16.7) | 3 | 2 (25.0) |
| **Skin and subcutaneous tissue disorders** | **1** | **1 (16.7)** | **-** | **-** | **-** | **-** | **-** | **-** | **4** | **4 (50.0)** |
| *Dermatitis contact* | - | - | - | - | - | - | - | - | 3 | 3 (37.5) |

**Abbreviations: AE = adverse event; BID = bis in diem/twice a day; n= number; QD = quaque die/once a day; SAE = serious adverse event.**

**Table S2: Effects of NMD670 versus placebo on handgrip release profile, for single- and multiple-ascending doses. The table shows post-dose estimated means for placebo and NMD670; and the estimated difference, with 95% confidence interval of the estimated mean, and *p*-value.**

| | | **Single-ascending dose study** | | | | | | | **Multiple-ascending dose study** | | | |
|---|---|---|---|---|---|---|---|---|---|---|---|---|
| | | Cohort 1 | | | Cohort 2 | | | Cohort 3 | Cohort 1 | Cohort 2 | Cohort 3 | Cohort 4 |
| **Dose** | | NMD670 50mg | NMD670 100mg | NMD670 200mg | NMD670 400mg | NMD670 800mg | NMD670 1200mg | NMD670 1200mg | NMD670 200mg QD | NMD670 400mg QD | NMD670 600mg QD | NMD670 400mg BID |
| **Grip Maximum Voluntary Contractions (kg)** | *ED* | -4.97 | -2.03 | -0.213 | 0.284 | -0.483 | 2.95 | -1.22 | 4.13 | 1.81 | -2.36 | 4.43 |
| | *95% CI* | (-7.52, -2.41) | (-4.62, 0.556) | (-2.79, 2.37) | (-2.10, 2.66) | (-2.87, 1.90) | (0.53, 5.37) | (-4.23, 1.80) | (-0.903, 9.15) | (-3.17, 6.79) | (-7.43, 2.70) | (-0.460, 9.33) |
| | *p-value* | **p<0.001** | p=0.123 | p=0.871 | p=0.814 | p=0.690 | **p=0.0171** | p=0.419 | p=0.104 | p=0.462 | p=0.346 | p=0.0739 |
| **Grip Peak Force (kg)** | *ED* | -4.19 | -1.95 | -1.05 | 0.882 | -1.29 | 2.27 | -0.105 | 4.86 | 1.50 | -2.10 | 4.21 |
| | *95% CI* | (-6.43, -1.94) | (-4.23, 0.334) | (-3.33, 1.23) | (-1.24, 3.00) | (-3.33, 0.753) | (0.129, 4.41) | (-2.74, 2.53) | (0.237, 9.47) | (-3.18, 6.18) | (-6.81, 2.61) | (-0.340, 8.76) |
| | *p-value* | **p<0.001** | p=0.0938 | p=0.366 | p=0.412 | p=0.214 | **p=0.0378** | p=0.936 | **p=0.0401** | p=0.515 | p=0.367 | p=0.0683 |
| **Grip Release Time 90% - 5% of the MVC (ms)** | *ED* | -8.74 | 13.4 | -8.66 | -2.21 | -18.9 | 13.8 | 11.1 | 0.26 | -9.54 | -19.6 | -33.4 |
| | *95% CI* | (-55.5, 38.0) | (-34.7, 61.5) | (-56.7, 39.4) | (-46.4, 41.9) | ( -58.9, 21.0) | (-30.7, 58.3) | (-17.6, 39.8) | (-61.0, 61.5) | (-68.9, 49.9) | (-78.8, 39.6) | (-92.6, 25.8) |
| | *p-value* | p=0.713 | p=0.584 | p=0.722 | p=0.922 | p=0.351 | p=0.541 | p=0.439 | p=0.993 | p=0.743 | p=0.501 | p=0.255 |
| **Grip Release Time 90% - 50% of the MVC (ms)** | *ED* | -2.74 | 9.77 | 4.54 | 11.1 | -4.6 | 0.3 | -0.08 | 5.65 | -5.76 | 0.2 | -13.3 |
| | *95% CI* | (-19.4, 13.9) | (-7.00, 26.5) | (-11.9, 21.0) | (-4.58, 26.8) | (-18.0, 8.83) | (-15.5, 16.0) | (-20.6, 20.4) | (-17.9, 29.2) | (-29.3, 17.8) | (-22.9, 23.3) | (-36.3, 9.79) |
| | *p-value* | p=0.746 | p=0.252 | p=0.586 | p=0.164 | p=0.500 | p=0.971 | p=0.994 | p=0.624 | p=0.617 | p=0.986 | p=0.245 |

Abbreviations: ED = estimated difference; CI = confidence interval.

**Table S3: Evaluation of dose proportionality for pharmacokinetic parameters $AUC_{inf}$ and $C_{max}$. Dose proportionality can be confirmed when the 90% confidence interval (90% CI) of the slope of the regression falls within the critical confidence interval (CI).**

| | | | **90% CI** | | **Critical CI** | |
|---|---|---|---|---|---|---|
| | *P-value* | *Slope* | *Lower* | *Upper* | *Lower* | *Upper* |
| $AUC_{inf}$ | <.0001 | 1.18 | 1.09 | 1.28 | 0.91 | 1.09 |
| $C_{max}$ | <.0001 | 1.05 | 0.977 | 1.12 | 0.93 | 1.07 |

**Abbreviations: $AUC_{inf}$ = area under the concentration time curve from time 0 to infinity; $C_{max}$ = maximum concentration.**

**Table S4: Evaluation of food and gender effects on pharmacokinetic parameters $AUC_{inf}$ and $C_{max}$. To explore bioequivalence the back-transformed 90% confidence interval (CI) around the difference on log scale was compared with the 80.00% to 125.00% criteria.**

| | | | | **90% CI** | |
|---|---|---|---|---|---|
| | *Contrast* | *Ratio of Geometric LSMs* | *P-value* | *Lower* | *Upper* |
| $AUC_{inf}$ | Fed - Fasted | 0.95 | 0.218 | 0.87 | 1.02 |
| $C_{max}$ | Fed - Fasted | 1.08 | 0.705 | 0.73 | 1.61 |
| $AUC_{inf}$ | Female - Male | 1.33 | 0.0261 | 1.09 | 1.62 |
| $C_{max}$ | Female - Male | 1.30 | 0.229 | 0.9 | 1.87 |

**Abbreviations: $AUC_{inf}$ = area under the concentration time curve from time 0 to infinity; $C_{max}$ = maximum concentration; LSM = least square mean.**

**Table S5: Pharmacokinetic parameters of NMD670 in plasma following administration of all multiple ascending dose levels.**

| | | **200 mg QD** | | **400 mg QD** | | **600 mg QD** | | **400 mg BID** | |
|---|---|---|---|---|---|---|---|---|---|
| | | **Day 1** | **Day 10** | **Day 1** | **Day 10** | **Day 1** | **Day 10** | **Day 1** | **Day 10** |
| | | **N=6** | **N=5** | **N=6** | **N=6** | **N=6** | **N=6** | **N=6** | **N=6** |
| $C_{max}$ (ng/mL) | *Mean* | 9638.3 | 8916.0 | 24883.3 | 19583.3 | 27033.3 | 26200.0 | 28466.7 | 31250.0 |
| | *SD* | 3765.42 | 1548.41 | 4997.37 | 3142.88 | 6185.04 | 2041.57 | 4048.04 | 9336.11 |
| | *Min-max* | 2880-14000 | 7250-10700 | 18300-30300 | 14300-23600 | 19900-37400 | 23700-29000 | 22900-32800 | 22700-44000 |
| $T_{max}$ (h) | *Median* | 3.50 | 3.00 | 2.05 | 2.50 | 3.00 | 2.50 | 8.00 | 8.01 |
| | *Min-max* | 1.00-12.00 | 2.00-4.03 | 1.23-4.00 | 1.00-4.18 | 2.00-4.20 | 2.00-4.00 | 3.00-9.00 | 4.00-9.00 |
| $AUC_{inf}$ (h*ng/mL) | *Mean* | 46465.6 | - | 112764.1 | - | 147798.0 | - | 204183.0 | - |
| | *SD* | 6128.30 | - | 18471.09 | - | 55860.44 | - | 20797.79 | - |
| | *Min-max* | 42678-57353 | - | 84371-134362 | - | 95437-250496 | - | 172899-224666 | - |
| $AUC_{24h}$ (h*ng/mL) | *Mean* | 42743.3 | 40999.5 | 108527.2 | 85912.5 | 143686.2 | 124160.4 | 194549.3 | 185804.6 |
| | *SD* | 7360.55 | 6889.95 | 17069.71 | 16141.72 | 53390.79 | 34757.93 | 20638.84 | 29399.33 |
| | *Min-max* | 31575-54798 | 33292-49972 | 81128-127997 | 67153-112989 | 93570-241837 | 93278-189060 | 168641-219439 | 153908-225043 |
| CL/F (L/h) | *Mean* | - | 4.991 | - | 4.786 | - | 5.101 | - | 4.395 |
| | *SD* | - | 0.8418 | - | 0.8460 | - | 1.1806 | - | 0.6795 |
| | *Min-max* | - | 4.00-6.01 | - | 3.54-5.96 | - | 3.17-6.43 | - | 3.55-5.20 |
| $t_{1/2}$ (h) | *Mean* | 5.344 | 5.145 | 5.662 | 4.820 | 4.672 | 5.373 | 3.701 | 5.103 |
| | *SD* | 0.9713 | 0.7541 | 0.6883 | 0.6353 | 0.1925 | 1.9476 | 0.1574 | 1.2619 |
| | *Min-max* | 3.97-6.67 | 4.48-6.23 | 4.96-6.73 | 3.92-5.65 | 4.44-4.88 | 3.96-8.93 | 3.52-3.93 | 3.18-6.12 |
| $V_z/F$ (L) | *Mean* | - | 37.20 | - | 33.54 | - | 38.08 | - | 31.65 |
| | *SD* | - | 9.745 | - | 9.131 | - | 12.079 | - | 6.497 |
| | *Min-max* | - | 29.7-54.0 | - | 25.4-48.6 | - | 28.8-61.7 | - | 22.4-41.2 |
| Rac(AUC) | *Mean* | - | 0.918 | - | 0.798 | - | 0.887 | - | 0.955 |
| | *SD* | - | 0.1681 | - | 0.1280 | - | 0.0898 | - | 0.1077 |
| | *Min-max* | - | 0.77-1.18 | - | 0.62-0.95 | - | 0.78-1.00 | - | 0.80-1.12 |
| Rac($C_{max}$) | *Mean* | - | 0.818 | - | 0.811 | - | 1.011 | - | 1.113 |
| | *SD* | - | 0.1134 | - | 0.1994 | - | 0.2408 | - | 0.3605 |
| | *Min-max* | - | 0.68-0.96 | - | 0.62-1.14 | - | 0.73-1.37 | - | 0.76-1.72 |

**Abbreviations: AUC = area under the concentration time curve; $AUC_{inf}$ = area under the concentration-time curve from time zero to infinity; $AUC_{24h}$ = area under the concentration time curve from time zero to 24 hours; BID = bis in diem/twice a day; CL/F = apparent total clearance following extravascular administration; $C_{max}$ = maximum concentration; max = maximum; Min = minimum; N = number of participants in PK population; PK = pharmacokinetic; QD = quaque die/once a day; Rac =**

**drug accumulation ratio; SD = standard deviation; $T_{max}$ = time to maximum concentration; $t_{1/2}$ = terminal elimination half-life; Vz/F = apparent volume of distribution during the terminal elimination phase after extravascular administration.**
**Concentrations below limit of quantification (25 ng/mL) were set to 0.**
**For several dose levels, the N for certain parameters is smaller than the number listed at the top of the column.**

**Table S6 Effects of NMD670 versus placebo on muscle velocity recovery cycles, in single- and multiple-ascending doses. The table shows post-dose estimated means for placebo and NMD670; and the estimated difference, with 95% confidence interval of the estimated mean, and *p*-value.**

| | | Single-ascending dose study | | | | | | Multiple-ascending dose study | | | |
|---|---|---|---|---|---|---|---|---|---|---|---|
| | | Cohort 1 | | | Cohort 2 | | | Cohort 1 | Cohort 2 | Cohort 3 | Cohort 4 |
| | | NMD670 50mg | NMD670 100mg | NMD670 200mg | NMD670 400mg | NMD670 800mg | NMD670 1200mg | NMD670 200mg QD | NMD670 400mg QD | NMD670 600mg QD | NMD670 400mg BID |
| Latency (ms) | *ED* | 0.357 | -0.207 | -0.166 | -0.209 | 0.165 | 0.14 | 0.232 | 0.07 | 0.231 | 0.253 |
| | *95% CI* | (-0.171, 0.884) | (-0.644, 0.229) | (-0.610, 0.278) | (-0.656, 0.237) | (-0.296, 0.625) | (-0.299, 0.579) | (-0.350, 0.815) | (-0.497, 0.636) | (-0.345, 0.808) | (-0.314, 0.821) |
| | *p-value* | p=0.179 | p=0.342 | p=0.454 | p=0.349 | p=0.474 | p=0.522 | p=0.417 | p=0.801 | p=0.416 | p=0.365 |
| Relative refractory period (RRP) (ms) | *ED* | 0.131 | 0.221 | 0.443 | -0.056 | 0.231 | 0.16 | -0.084 | -0.541 | -0.08 | -0.238 |
| | *95% CI* | (-0.450, 0.713) | (-0.276, 0.719) | (-0.043, 0.929) | (-0.563, 0.451) | (-0.258, 0.721) | (-0.366, 0.686) | (-0.708, 0.540) | (-1.13, 0.047) | (-0.662, 0.501) | (-0.875, 0.400) |
| | *p-value* | p=0.649 | p=0.372 | p=0.0726 | p=0.824 | p=0.344 | p=0.541 | p=0.783 | p=0.0698 | p=0.778 | p=0.449 |
| Early supernormality (ESN) (%) | *ED* | 0.299 | -0.138 | -2.09 | 0.005 | -0.901 | -0.387 | -0.036 | 2.45 | -0.052 | -0.543 |
| | *95% CI* | (-2.80, 3.40) | (-2.76, 2.49) | (-4.74, 0.551) | (-2.67, 2.68) | (-3.53, 1.73) | (-3.06, 2.28) | (-3.52, 3.45) | (-0.980, 5.89) | (-3.54, 3.43) | (-4.08, 2.99) |
| | *p-value* | p=0.846 | p=0.916 | p=0.117 | p=0.997 | p=0.492 | p=0.771 | p=0.984 | p=0.154 | p=0.976 | p=0.755 |
| ISI corresponding to ESN (ESN@) (ms) | *ED* | 0.598 | 0.443 | 1.75 | -0.909 | 1.12 | 1.01 | 0.446 | -0.88 | 0.602 | -0.236 |
| | *95% CI* | (-1.19, 2.39) | (-1.06, 1.95) | ( 0.24, 3.26) | (-2.51, 0.697) | (-0.397, 2.65) | (-0.497, 2.53) | (-1.04, 1.93) | (-2.29, 0.532) | (-0.873, 2.08) | (-1.81, 1.34) |
| | *p-value* | p=0.503 | p=0.554 | **p=0.0242** | p=0.258 | p=0.142 | p=0.181 | p=0.541 | p=0.211 | p=0.409 | p=0.760 |
| Early supernormality after 5 CS (5ESN) (%) | *ED* | 0.337 | 1.11 | -1.53 | -0.766 | -0.964 | -0.244 | 0.167 | 3.20 | 0.128 | 0.332 |
| | *95% CI* | (-2.38, 3.05) | (-1.21, 3.43) | (-3.84, 0.787) | (-3.11, 1.57) | (-3.28, 1.35) | (-2.60, 2.11) | (-2.84, 3.17) | ( 0.270, 6.13) | (-2.82, 3.08) | (-2.75, 3.41) |
| | *p-value* | p=0.803 | p=0.339 | p=0.190 | p=0.511 | p=0.405 | p=0.835 | p=0.910 | **p=0.0336** | p=0.930 | p=0.826 |
| Supernormality at 20 ms (SN20) (%) | *ED* | -0.05 | 0.867 | -0.189 | -0.429 | 0.851 | 0.759 | - | - | - | - |
| | *95% CI* | (-1.73, 1.62) | (-0.72, 2.45) | (-1.85, 1.47) | (-2.09, 1.23) | (-0.71, 2.42) | (-0.87, 2.39) | - | - | - | - |
| | *p-value* | p=0.952 | p=0.275 | p=0.820 | p=0.605 | p=0.278 | p=0.351 | - | - | - | - |

| | | | | | | | | | | | |
|---|---|---|---|---|---|---|---|---|---|---|---|
| Late supernormality (LSN) (%) | *ED* | 0.15 | 0.327 | 0.53 | -0.405 | 0.38 | -0.137 | -0.875 | -0.384 | -0.463 | -0.576 |
| | *95% CI* | (-0.747, 1.05) | (-0.431, 1.09) | (-0.226, 1.29) | (-1.18, 0.369) | (-0.404, 1.16) | (-0.960, 0.687) | (-1.79, 0.044) | (-1.30, 0.527) | (-1.38, 0.451) | (-1.49, 0.337) |
| | *p-value* | p=0.735 | p=0.385 | p=0.162 | p=0.296 | p=0.332 | p=0.737 | p=0.0612 | p=0.393 | p=0.307 | p=0.206 |
| Late supernormality 2 - 1 CS (2XLSN) (%) | *ED* | -0.112 | 0.036 | -0.053 | -0.061 | 0.191 | -0.522 | -0.417 | 0.429 | 0.01 | 0.183 |
| | *95% CI* | (-0.661, 0.438) | (-0.438, 0.509) | (-0.525, 0.419) | (-0.527, 0.404) | (-0.277, 0.660) | (-1.02, -0.020) | (-0.962, 0.129) | (-0.109, 0.967) | (-0.536, 0.555) | (-0.364, 0.731) |
| | *p-value* | p=0.683 | p=0.879 | p=0.821 | p=0.791 | p=0.413 | **p=0.0421** | p=0.129 | p=0.113 | p=0.971 | p=0.496 |
| Late supernormality 5 - 1 CS (5XLSN) (%) | *ED* | -0.368 | 0.041 | -0.076 | -0.023 | 0.378 | -0.849 | -0.799 | 1.17 | -0.098 | 0.678 |
| | *95% CI* | (-1.51, 0.768) | (-0.933, 1.01) | (-1.07, 0.918) | (-0.986, 0.941) | (-0.587, 1.34) | (-1.89, 0.190) | (-1.86, 0.264) | ( 0.110, 2.22) | (-1.15, 0.950) | (-0.397, 1.75) |
| | *p-value* | p=0.516 | p=0.933 | p=0.877 | p=0.963 | p=0.433 | p=0.106 | p=0.134 | **p=0.0317** | p=0.849 | p=0.206 |
| Residual supernormality (RSN) (%) | *ED* | 0.014 | -0.048 | 0.14 | 0.21 | 0.095 | -0.01 | -0.007 | 0.035 | -0.129 | -0.099 |
| | *95% CI* | (-0.274, 0.301) | (-0.308, 0.211) | (-0.101, 0.381) | (-0.033, 0.454) | (-0.150, 0.341) | (-0.253, 0.234) | (-0.234, 0.220) | (-0.187, 0.257) | (-0.354, 0.095) | (-0.318, 0.120) |
| | *p-value* | p=0.924 | p=0.708 | p=0.246 | p=0.0879 | p=0.438 | p=0.936 | p=0.950 | p=0.747 | p=0.247 | p=0.361 |
| Residual supernormality 2 - 1 CS (2XRSN) (%) | *ED* | -0.068 | -0.07 | -0.242 | 0.046 | 0.122 | -0.035 | -0.363 | 0.311 | -0.031 | -0.088 |
| | *95% CI* | (-0.524, 0.388) | (-0.362, 0.221) | (-0.540, 0.056) | (-0.263, 0.356) | (-0.173, 0.418) | (-0.308, 0.239) | (-0.782, 0.056) | (-0.091, 0.713) | (-0.441, 0.378) | (-0.493, 0.317) |
| | *p-value* | p=0.764 | p=0.625 | p=0.108 | p=0.762 | p=0.406 | p=0.798 | p=0.0867 | p=0.124 | p=0.876 | p=0.659 |
| Residual supernormality 5 - 1 CS (5XRSN) (%) | *ED* | -0.188 | -0.493 | 0.088 | -0.047 | -0.028 | -0.482 | - | - | - | - |
| | *95% CI* | (-0.720, 0.343) | (-0.933, -0.052) | (-0.321, 0.497) | (-0.468, 0.374) | (-0.457, 0.400) | (-0.895, -0.069) | - | - | - | - |

| | | | | | | | | | | | |
|---|---|---|---|---|---|---|---|---|---|---|---|
| | *p-value* | p=0.476 | **p=0.0297** | p=0.664 | p=0.822 | p=0.895 | **p=0.0237** | - | - | - | - |
| Mean CMAP amplitude (mV) | *ED* | 0.07 | 0.128 | 0.043 | 0.06 | 0.073 | 0.137 | -0.073 | -0.082 | -0.052 | -0.1 |
| | *95% CI* | (-0.089, 0.230) | (-0.017, 0.274) | (-0.094, 0.179) | (-0.081, 0.200) | (-0.064, 0.209) | ( 0.000, 0.273) | (-0.161, 0.015) | (-0.164, 0.001) | (-0.138, 0.034) | (-0.189, -0.010) |
| | *p-value* | p=0.377 | p=0.0826 | p=0.534 | p=0.395 | p=0.288 | **p=0.0492** | p=0.101 | p=0.0532 | p=0.224 | **p=0.0300** |
| Lat (15Hz) first (%) | *ED* | -1.07 | -0.335 | 0.052 | -0.295 | -0.281 | 0.306 | 1.042 | -1.27 | -0.102 | 0.505 |
| | *95% CI* | (-2.90, 0.773) | (-2.04, 1.37) | (-1.65, 1.76) | (-2.01, 1.42) | (-2.01, 1.45) | (-1.40, 2.01) | (-1.01, 3.10) | (-3.23, 0.70) | (-2.14, 1.94) | (-1.48, 2.49) |
| | *p-value* | p=0.248 | p=0.693 | p=0.951 | p=0.729 | p=0.744 | p=0.718 | p=0.306 | p=0.196 | p=0.919 | p=0.603 |
| Lat (15Hz) last (%) | *ED* | 0.843 | -0.895 | 0.455 | 0.613 | -0.518 | 1.74 | 1.17 | -1.27 | 0.623 | 1.04 |
| | *95% CI* | (-1.69, 3.38) | (-3.09, 1.30) | (-1.73, 2.64) | (-1.77, 2.99) | (-2.70, 1.66) | (-0.43, 3.92) | (-1.32, 3.66) | (-3.75, 1.21) | (-1.81, 3.06) | (-1.37, 3.45) |
| | *p-value* | p=0.505 | p=0.413 | p=0.676 | p=0.605 | p=0.633 | p=0.113 | p=0.341 | p=0.300 | p=0.601 | p=0.381 |
| Peak (15Hz) last (%) | *ED* | -9.34 | 8.50 | -5.93 | 7.29 | -22.9 | -7.34 | 7.28 | 3.84 | -1.88 | -8.69 |
| | *95% CI* | (-44.3, 25.7) | (-19.8, 36.8) | (-33.5, 21.7) | (-23.3, 37.9) | (-51.0, 5.28) | (-34.9, 20.3) | (-11.1, 25.6) | (-14.0, 21.7) | (-20.1, 16.3) | (-26.0, 8.66) |
| | *p-value* | p=0.593 | p=0.546 | p=0.665 | p=0.632 | p=0.108 | p=0.593 | p=0.422 | p=0.661 | p=0.833 | p=0.311 |
| Peak (15Hz) first (%) | *ED* | 5.90 | 4.18 | 4.95 | 5.47 | -4.13 | 3.33 | 0.618 | 6.61 | 3.59 | -1.64 |
| | *95% CI* | (-11.0, 22.8) | (-11.7, 20.0) | (-10.5, 20.4) | (-10.2, 21.2) | (-19.8, 11.5) | (-12.1, 18.8) | (-6.60, 7.83) | (-0.27, 13.5) | (-3.39, 10.6) | (-8.57, 5.28) |
| | *p-value* | p=0.484 | p=0.597 | p=0.520 | p=0.485 | p=0.596 | p=0.665 | p=0.861 | p=0.0588 | p=0.297 | p=0.626 |
| Lat (30Hz) first (%) | *ED* | 0.162 | -0.112 | 0.24 | 0 | -0.428 | 2.56 | 0.27 | -0.835 | -0.545 | -1.46 |
| | *95% CI* | (-2.42, 2.74) | (-2.54, 2.31) | (-2.18, 2.66) | (-2.40, 2.40) | (-2.94, 2.09) | ( 0.124, 5.00) | (-2.46, 3.00) | (-3.55, 1.88) | (-3.22, 2.13) | (-3.97, 1.05) |
| | *p-value* | p=0.900 | p=0.926 | p=0.842 | p=1.00 | p=0.733 | **p=0.0399** | p=0.841 | p=0.532 | p=0.679 | p=0.242 |
| Peak (30Hz) first (%) | *ED* | 8.41 | 18.19 | 6.11 | 11.0 | -3.80 | 11.64 | 0.747 | 8.69 | 13.4 | -5.52 |
| | *95% CI* | (-13.1, 29.9) | (-2.35, 38.7) | (-13.7, 25.9) | (-8.99, 30.9) | (-23.9, 16.3) | (-8.13, 31.4) | (-12.0, 13.5) | (-4.17, 21.5) | ( 0.256, 26.6) | (-17.8, 6.76) |
| | *p-value* | p=0.434 | p=0.0810 | p=0.535 | p=0.273 | p=0.704 | p=0.241 | p=0.904 | p=0.174 | **p=0.0461** | p=0.361 |
| | *ED* | -4.22 | 20.4 | 10.5 | 12.1 | -18.8 | -8.68 | 15.0 | 8.97 | 6.78 | -12.6 |

| | | | | | | | | | | | |
|---|---|---|---|---|---|---|---|---|---|---|---|
| Peak (30Hz) last (%) | *95% CI* | (-36.1, 27.7) | (-9.09, 49.9) | (-18.7, 39.7) | (-18.0, 42.2) | (-48.2, 10.5) | (-37.8, 20.5) | (-10.7, 40.7) | (-17.7, 35.6) | (-20.5, 34.1) | (-37.1, 11.9) |
| | *p-value* | p=0.791 | p=0.170 | p=0.471 | p=0.422 | p=0.202 | p=0.550 | p=0.240 | p=0.493 | p=0.612 | p=0.300 |
| Peak (30-15Hz) first (%) | *ED* | 1.38 | 14.4 | 1.98 | 5.34 | -0.641 | 6.54 | -3.02 | -1.67 | 5.73 | -6.22 |
| | *95% CI* | (-9.80, 12.6) | ( 3.88, 25.0) | (-8.41, 12.4) | (-5.04, 15.7) | (-11.1, 9.81) | (-3.84, 16.9) | (-14.2, 8.19) | (-13.0, 9.68) | (-6.28, 17.7) | (-17.7, 5.23) |
| | *p-value* | p=0.804 | **p=0.0086** | p=0.702 | p=0.305 | p=0.902 | p=0.210 | p=0.586 | p=0.764 | p=0.337 | p=0.275 |
| Lat (30Hz+30s) (%) | *ED* | -0.199 | 1.17 | 0.573 | 0.994 | 0.069 | 0.935 | -1.13 | -1.79 | -0.07 | -1.83 |
| | *95% CI* | (-1.76, 1.37) | (-0.239, 2.58) | (-0.816, 1.96) | (-0.474, 2.46) | (-1.43, 1.57) | (-0.467, 2.34) | (-2.86, 0.603) | (-3.38, -0.196) | (-1.76, 1.62) | (-3.58, -0.080) |
| | *p-value* | p=0.798 | p=0.101 | p=0.407 | p=0.178 | p=0.926 | p=0.184 | p=0.192 | **p=0.0294** | p=0.933 | **p=0.0411** |
| Peak (30Hz+30s) (%) | *ED* | 13.1 | 7.75 | 4.97 | 11.7 | 5.37 | 3.44 | -2.48 | -1.70 | 13.4 | -6.13 |
| | *95% CI* | (0.877, 25.3) | (-3.88, 19.4) | (-6.07, 16.0) | (-0.420, 23.8) | (-5.96, 16.7) | (-7.63, 14.5) | (-11.5, 6.49) | (-10.2, 6.81) | ( 4.67, 22.2) | (-14.7, 2.49) |
| | *p-value* | **p=0.0364** | p=0.185 | p=0.366 | p=0.0580 | p=0.342 | p=0.531 | p=0.574 | p=0.682 | **p=0.0042** | p=0.155 |
| LatMinFirst (%) | *ED* | 0.02 | -0.72 | 0.81 | 0.02 | 0.14 | 0.55 | - | - | - | - |
| | *95% CI* | ( -2.01, 2.05) | ( -2.60, 1.17) | ( -1.08, 2.70) | ( -1.87, 1.90) | ( -1.79, 2.07) | ( -1.34, 2.43) | - | - | - | - |
| | *p-value* | p=0.984 | p=0.447 | p=0.390 | p=0.984 | p=0.884 | p=0.560 | - | - | - | - |
| LatMinLast (%) | *ED* | 0.79 | -0.68 | 0.48 | 0.31 | -0.07 | 1.64 | - | - | - | - |
| | *95% CI* | ( -1.90, 3.48) | ( -3.20, 1.83) | ( -2.03, 2.99) | ( -2.21, 2.83) | ( -2.59, 2.44) | ( -0.86, 4.14) | - | - | - | - |
| | *p-value* | p=0.556 | p=0.586 | p=0.700 | p=0.806 | p=0.953 | p=0.193 | - | - | - | - |
| FreqLatMinFirst (Hz) | *ED* | -2.01 | 0.26 | -0.25 | -2.15 | -0.25 | -4.8 | - | - | - | - |
| | *95% CI* | ( -6.93, 2.90) | ( -4.32, 4.83) | ( -4.83, 4.34) | ( -6.73, 2.42) | ( -4.85, 4.36) | ( -9.38, -0.22) | - | - | - | - |
| | *p-value* | p=0.413 | p=0.911 | p=0.914 | p=0.347 | p=0.915 | p=0.0404 | - | - | - | - |
| FreqLatMinLast (Hz) | *ED* | -1.7 | -1.09 | -0.75 | -0.77 | -0.31 | 0.67 | - | - | - | - |
| | *95% CI* | ( -5.20, 1.80) | ( -4.33, 2.14) | ( -4.00, 2.50) | ( -4.12, 2.58) | ( -3.61, 2.99) | ( -2.60, 3.93) | - | - | - | - |

| | | | | | | | | | | | |
|---|---|---|---|---|---|---|---|---|---|---|---|
| | *p-value* | p=0.332 | p=0.498 | p=0.643 | p=0.644 | p=0.851 | p=0.681 | - | - | - | - |

**Abbreviations: ED = estimated difference; CI = confidence interval.**
